# Decoupling Accuracy and Explainability: Machine Learning Strategies for HbA1c Prediction and Biomarker Discovery in Blood FTIR Spectroscopy

**DOI:** 10.64898/2026.01.26.26344831

**Authors:** Mykola Melnychenko, Tatiana Makhnii, Konstantin Midlovets, Bogdan Dmyterchuk, Dmytro Krasnienkov

## Abstract

Glycated hemoglobin (HbA1c) is a central biomarker for long-term glycemic control and diabetes management, traditionally quantified using laboratory-intensive chromatographic or immunochemical assays. As the global burden of diabetes continues to rise, there is growing interest in alternative, scalable approaches capable of rapid biochemical assessment. Fourier-transform infrared (FTIR) spectroscopy offers a reagent-free method that captures molecular signatures of protein glycation, but translating complex spectra into clinically interpretable HbA1c values requires robust analytical frameworks.

Here, we present a complementary multi-model strategy for predicting HbA1c from FTIR spectra of whole blood. Using 685 blood samples with matched reference HbA1c measurements, we evaluated three analytically distinct yet synergistic approaches: partial least squares regression (PLSR), peak-resolved curve fitting based on pseudo-Voigt functions combined with H2O AutoML, and a convolutional neural network (CNN). PLSR and CNN models were trained on biologically informative spectral regions (800–1800 cm⁻¹ and 2800–3400 cm⁻¹), while curve fitting focused on the fingerprint region (1000–1720 cm⁻¹) to extract interpretable biochemical parameters.

PLSR achieved the highest predictive accuracy (R² = 0.76), closely followed by the CNN (R² = 0.73), reflecting their ability to capture global linear and nonlinear spectral relationships. Although curve fitting yielded lower predictive performance (R² = 0.59), its peak-level decomposition enabled mechanistic interpretation of glycation-related changes. Explainable AI analysis using SHAP identified lipid- and protein-associated vibrations, carbohydrate-linked glycation bands, and amide-region structural features as key contributors to HbA1c prediction.

Rather than treating these approaches as competing alternatives, our results demonstrate that their integration provides a more informative framework than any single model alone. By combining predictive performance with biochemical interpretability, this multi-model FTIR strategy highlights a scalable and mechanistically grounded pathway toward non-invasive HbA1c assessment and broader metabolic screening in diabetes monitoring. The code for this study is freely available at https://github.com/MelnychenkoM/ftir-hba1c-prediction.

## Introduction

Glycated hemoglobin (HbA1c) has been the cornerstone of diabetes diagnosis and monitoring since its adoption in 1979, offering a robust measure of long-term glycemic control that reflects average blood glucose levels over the 120-day lifespan of red blood cells [1, 2, 3, 4]. Its clinical importance continues to grow in the context of the worldwide increase in diabetes incidence. The global prevalence of diabetes has doubled since 1990, affecting 14% of adults by 2022, and is projected to reach 1.3 billion individuals by 2050 [5, 6]. Diabetes now accounts for more than 1.6 million deaths annually, nearly half of which occur prematurely, and is associated with a spectrum of severe complications, including nephropathy, retinopathy, neuropathy, and cardiovascular disease. Conventional HbA1c assays rely on high-performance liquid chromatography and immunoassays [7, 8]. While accurate, these methods require time, specialized reagents, and instrumentation, limiting scalability in low-resource settings. By contrast, Fourier-transform infrared (FTIR) spectroscopy is a reagent-free, rapid, and minimally invasive approach that sensitively captures molecular alterations associated with protein glycation [9, 10, 11, 12]. Modifications alter vibrational signatures across carbohydrate, protein, and amide regions, producing spectral fingerprints that can be exploited for quantitative biomarker development [16, 17]. Glycation proceeds via the Maillard reaction, in which glucose irreversibly modifies amino groups of hemoglobin, particularly at β-Val1, β-Lys66, and α-Lys61, forming stable ketoamine adducts [13, 14, 15].

However, interpreting complex FTIR spectra requires advanced computational methods. Classical approaches such as Principal Component Analysis (PCA), Principal Component Regression (PCR), and Partial Least Squares Regression (PLSR) have been widely applied, but they face limitations: PCA and PCR prioritize variance rather than biochemical relevance, while PLSR, though supervised, may be constrained by overlapping spectral features [18, 19]. Curve fitting using pseudo-Voigt profiles offers improved peak resolution and interpretability by decomposing broad signals into biochemically meaningful components [20–22]. Meanwhile, convolutional neural networks (CNNs) have shown remarkable success in automatically identifying functional groups and highlighting discriminative spectral regions without extensive preprocessing [23–26].

FTIR spectroscopy holds promise as a practical tool for clinical monitoring of long-term blood glucose levels and for the diagnosis of diabetes. Its simplicity, rapid acquisition, and minimal sample preparation make it well suited for preliminary screening workflows, including potential applications in public settings such as pharmacies, outpatient clinics, or other community-based facilities.

Importantly, the approach can be directly adapted to yield clinically interpretable values of HbA1c. Once a predictive model is calibrated against reference laboratory methods (e.g., HPLC or immunoassay), FTIR spectra of patient blood can be processed to output HbA1c estimates in the same standardized units (%), allowing straightforward integration into existing diagnostic frameworks. This eliminates the need for complex biochemical assays while still providing results that are compatible with international guidelines for diabetes monitoring.

To evaluate diagnostic performance, this study compares Partial Least Squares Regression (PLSR), Convolutional Neural Networks (CNN), and curve fitting – contrasting the interpretation of specific peaks with the predictive power of deep learning to achieve robust and explainable FTIR-based classification.

Despite these advances, few studies have compared or integrated PLSR, peak-resolved curve fitting, and CNN-based models for HbA1c prediction from FTIR blood spectra [27]. Because these approaches offer complementary strengths - interpretability, mechanistic insight, and nonlinear pattern recognition - their combined application may enhance spectral analysis.

Here, we evaluate PLSR, curve fitting integrated with H2O AutoML, and CNN models on 685 blood samples with reference HbA1c values. Although initially we aimed to explore these models as alternative strategies, these approaches are evaluated here as potentially complementary in HbA1c prediction.

## Materials and Methods

### Test Subjects

Informed consent was obtained from all participants before the collection of blood Fsamples. A total of 685 samples were collected using 4 mL EDTA-containing vacutainers and analyzed for HbA1c with high-performance ionic liquid chromatography (Table 1).

For FTIR spectra acquisition, 100 μL of whole blood from every participant was lyophilized at room temperature for 24 hours. The samples were subsequently analyzed using a Nicolet iS50 FTIR spectrometer equipped with an ATR accessory, allowing for single-reflection measurements. Each spectrum was recorded at a resolution of 4 cm⁻¹, with 32 scans per sample to ensure high spectral accuracy. High-performance ionic liquid chromatography was used as a reference method.

Six samples were identified as outliers and manually removed, leaving 679 samples for analysis. Additionally, six samples with HbA1c levels exceeding 14% were excluded due to their potential to skew the dataset, resulting in a final dataset of 673 samples. To evaluate the performance of different predictive models, 20% of the data was reserved for validation. Stratified sampling was employed based on a prior discretization into 8 bins according to HbA1c levels to ensure a representative distribution across the dataset (Fig. 2B).

**Table 1.** Study participant information and demographics.

| **Group** | **Number of participants** | **HbA1c range (%**<br>**concentration)** |
| --- | --- | --- |
| Healthy | 250 | 3.57–5.7 |
| Prediabetic | 78 | 5.72–6.5 |
| Diabetic | 345 | 6.58–14 |

A total of 685 blood samples were analyzed, comprising healthy (n = 250), prediabetic (n = 78), and diabetic (n = 345) cohorts. Examination of the FTIR spectra enabled the identification of absorption frequencies most relevant for differentiating among these groups.

### Spectral Acquisition

Samples were directly applied onto the ATR crystal. FTIR spectra were recorded in the range of 4000 to 400 cm⁻¹. Before starting the measurements, the ATR crystal was cleaned with ethanol 70%; and before each new sample, the ATR crystal was cleaned with ethanol 70%. A background spectrum was taken before measuring every new sample to account for environmental changes. The scheme of mathematical processing and statistical analysis of data is presented in Figure 3. Raw and preprocessed FTIR spectra are presented in the Supplementary Information Figure 1 and Figure 2.

**Figure 1.**
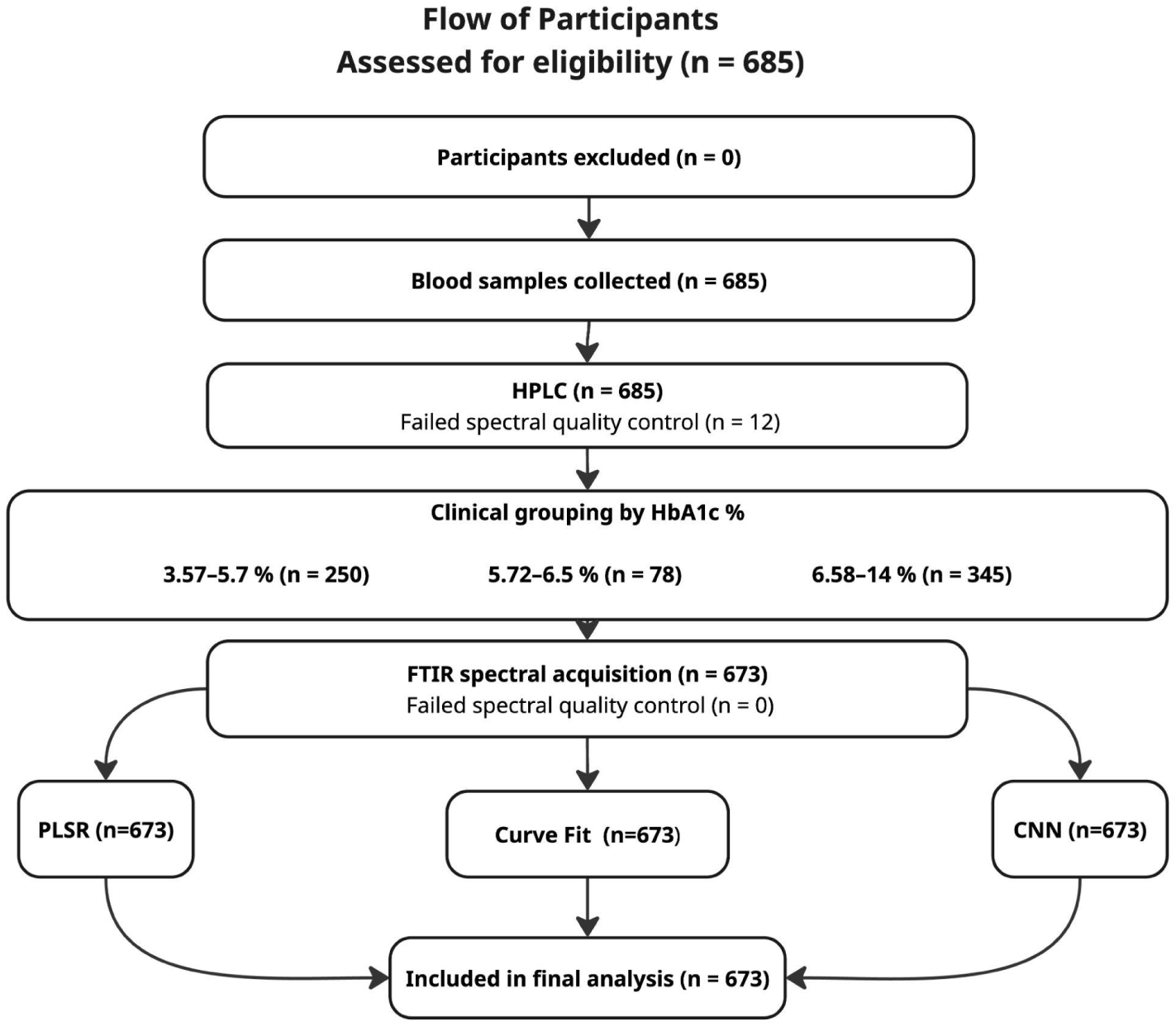
Flow of participants in the FTIR-based HbA1c study. A total of 685 participants were enrolled, with no exclusions. After HPLC reference analysis and quality control, 673 blood samples were retained for FTIR spectral acquisition. Spectra were grouped by clinical status (non-diabetic, prediabetic, diabetic) and analyzed using PLSR, curve-fitting, and CNN models to predict HbA1c levels.

**Figure 2.**
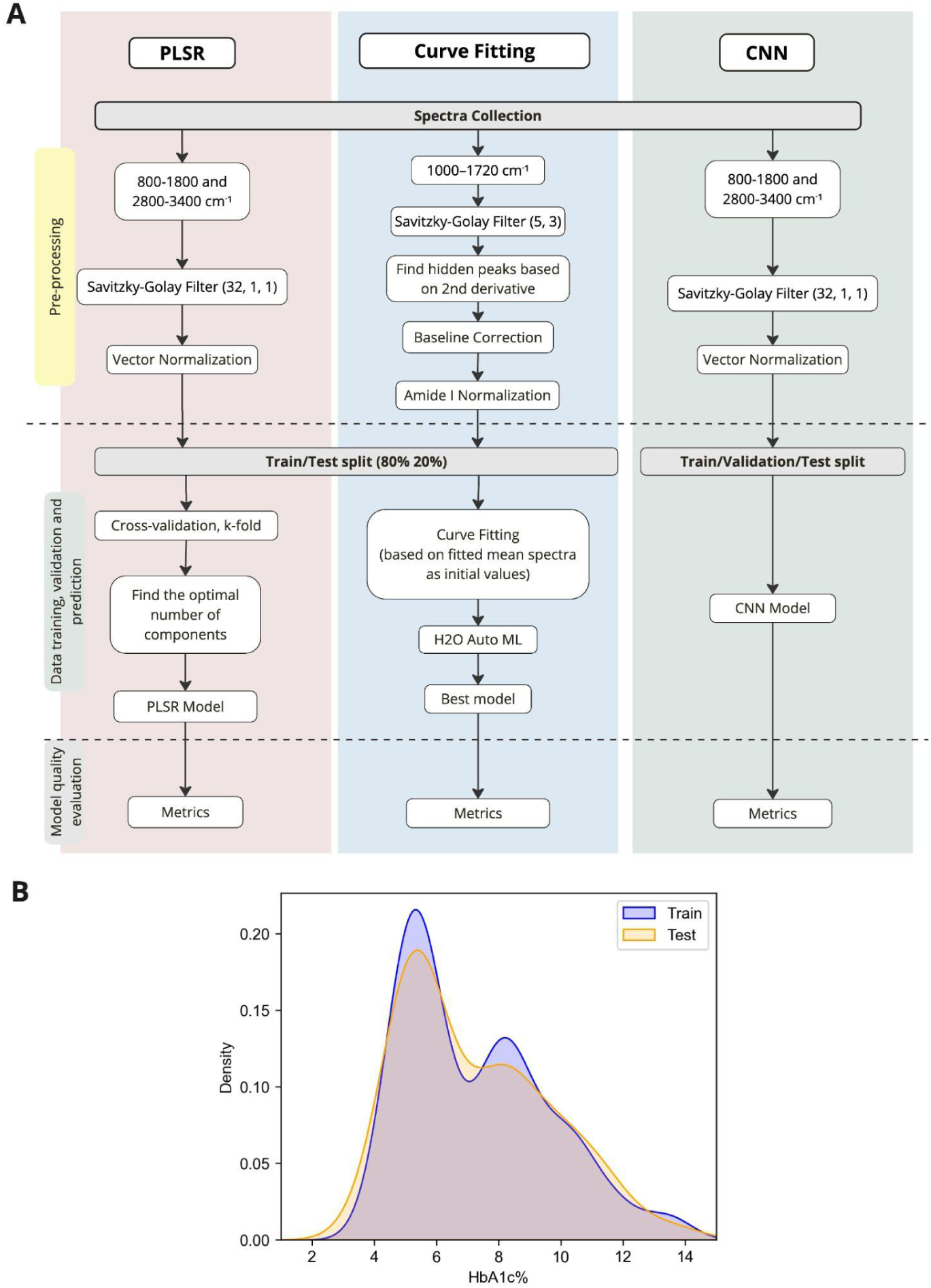
(A) Schematic representation of the computational pipeline for predicting HbA1c levels using three different approaches: PLSR, Curve Fitting with H2O AutoML, and CNNs. The workflow consists of spectral pre-processing, dataset splitting, model training, and evaluation. (B) Kernel density estimation (KDE) plot illustrating the distribution of HbA1c values in the training (blue) and test (orange) datasets, showing similar distributions, which ensures a representative data split.

**Figure 3.**
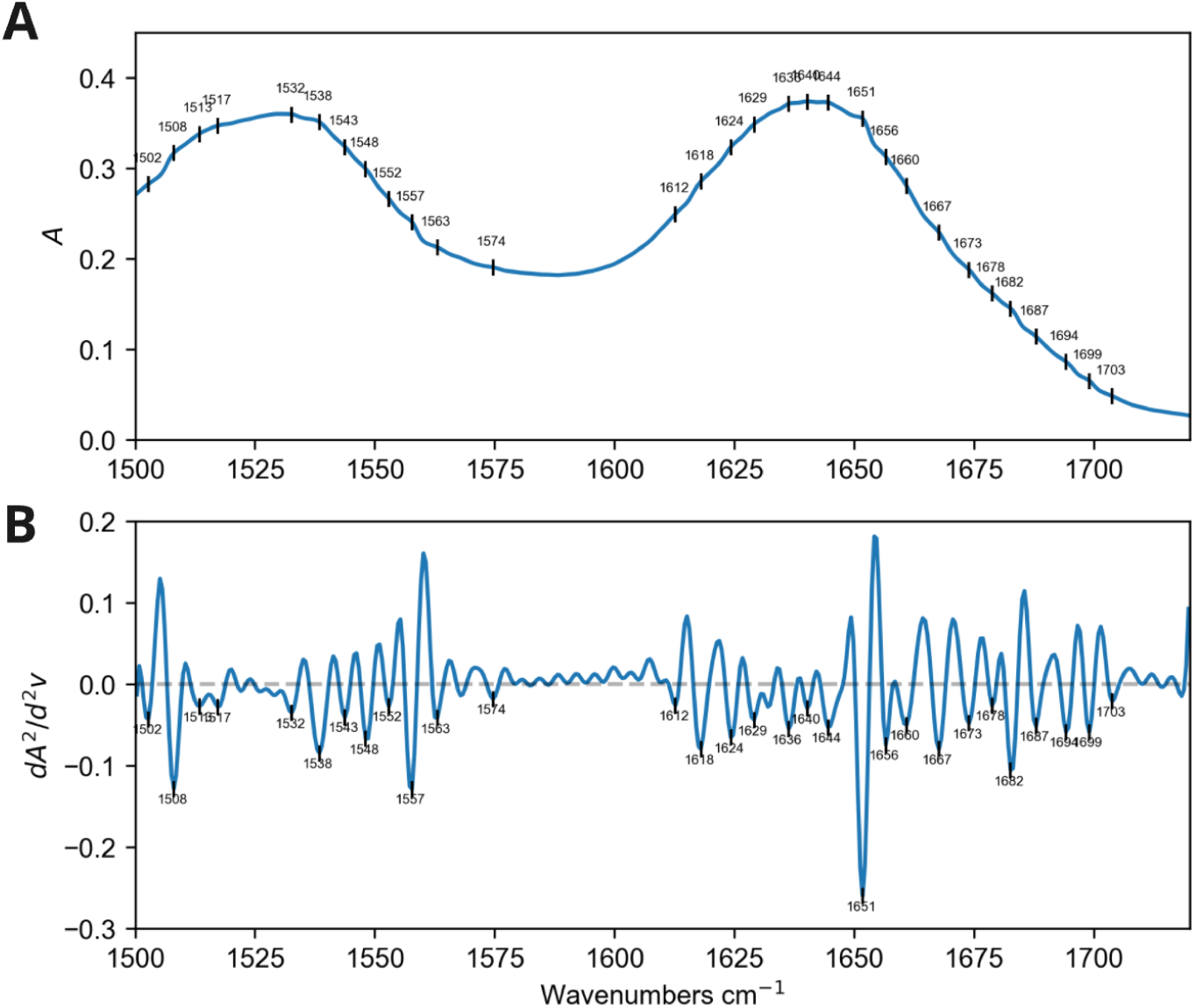
(A) the original absorbance spectrum, with detected peak positions indicated. Prominent bands are observed at 1502–1574 cm⁻¹ (amide II) and 1612–1703 cm⁻¹ (amide I), corresponding to protein secondary structure features; (B) second derivative and detected peaks on the individual spectrum. This approach makes it easier to distinguish peaks that overlap in the original spectrum. In the second derivative plot, the true peaks appear as clear minima, matching the positions of the bands seen in the raw spectrum and confirming their location within the amide regions (Only Amide I and Amide II regions are shown).

### PLSR

The PLSR model was trained using two spectral regions: 800–1800 cm⁻¹ and 2800–3400 cm⁻¹, as these contain the majority of biologically relevant information for blood samples. Pre-processing of the data involved applying a Savitzky-Golay filter (window length = 32, polynomial order = 2, first derivative) followed by vector normalization to enhance the quality of the spectra and reduce noise. The dataset was split into two sets: 80% of the samples were allocated to the training, while the remaining 20% were reserved for the testing (Figure 2A).

Due to the uneven distribution of glycated hemoglobin values, k-bin discretization with 8 bins and a uniform strategy was applied to stratify the data. The training dataset was utilized for model training, ensuring the model could capture the spectral patterns associated with HbA1c levels, while the test set was used to assess the model’s performance on unseen data, ensuring robust predictive capabilities.

The PLSR model was developed to predict HbA1c levels, with the dataset split into train and test sets. Cross-validation (CV) was employed to determine the optimal number of components in the PLSR model, which was then validated using an independent test set.

Selecting the optimal number of components in PLSR is crucial to avoid overfitting, especially in datasets with limited size. Cross-validation, typically k-fold (CV□), was employed to ensure model generalization. Training data were split into k subsets, with each subset serving as the validation set in turn. Larger k values (e.g., 10) maximize training data in smaller datasets, while smaller k values (e.g., 5) are computationally efficient for larger datasets. Predicted values from all folds were combined to compute the Root Mean Square Error of Cross-Validation (RMSECV). The model with the lowest RMSECV was deemed optimal.

To determine the optimal number of components, 10-fold cross-validation (CV_10_) was applied. The optimal number of components obtained from this process was then used to train and validate the model on the remaining test data, which comprised 20% of the dataset. PLSR was applied to the spectra collected from human blood samples in the 800 to 1800 cm⁻¹ and 2800 to 3400 cm⁻¹ regions.

### Curve Fitting

Three spectral regions were evaluated for HbA1c prediction with Curve Fitting. However, the 600–1000 cm⁻¹ and 2800–3400 cm⁻¹ regions, while potentially informative, were excluded due to excessive noise that obstructed model fitting. The 1000–1720 cm⁻¹ region was prioritized for further analysis, given its rich content of critical information on protein structures and carbohydrates, with differentiation enhancing the detection of concealed peaks. As illustrated in Figure 3A prominent bands are observed at 1502–1574 cm⁻¹ (amide II) and 1612–1703 cm⁻¹ (amide I), which correspond to protein secondary structure features. Figure 3B shows the corresponding second derivative spectrum with detected peaks, which improves resolution of overlapping signals.

Preprocessing for curve fitting involved spectral smoothing using a Savitzky–Golay filter with window size 5 and polynomial order 3 (Fig. 2A). The respective peaks in the second-derivative spectra were then detected, and only those appearing in at least 10% of all samples were retained for further analysis (Supplementary Information Figure 3). Next, rubberband baseline correction using the BoxSERS module (https://github.com/ALebrun-108/BoxSERS) was applied. The spectra were normalized based on the intensity of the Amide I peak to standardize comparisons.

The dataset was divided into two subsets: a training set (80%) and a testing set (20%). Also, k-bin discretization with 8 bins and a uniform strategy was applied. Next, the nonlinear curve fitting procedure was performed using the Trust Region Method implemented in the Python package JAXFit [28], which provides substantial speed improvements by leveraging GPU/TPU acceleration. The fitting function consists of the sum of all peaks, with each peak represented by a single pseudo-Voigt distribution. The pseudo-Voigt function approximates a Voigt profile by combining Gaussian and Lorentzian profiles.

It is important to note that the initialization of peak FWHM used a single value applied uniformly to all peaks. While this value was determined empirically, it does not account for individual peak characteristics. A more robust approach, recommended for future work, would involve calculating distinct initial values for each peak individually to improve algorithm convergence toward a biologically correct solution [122]. The detailed explanation of the curve fitting procedure can be found in Supplementary Information.

First, a primary model was fitted to the mean spectra of the dataset, and its parameters served as initial values for fitting each individual sample spectrum. This process demonstrated high accuracy, with discrepancies below 0.002 and R² values above 0.9999.

With the spectral data successfully quantified into a reliable set of parameters, the analysis transitioned to predictive modeling. These parameters were used to train H2O AutoML models via 5-fold cross-validation. Since the primary goal was to ensure interpretability of feature importance, ensemble and deep learning models were intentionally excluded. Finally, H2O AutoML’s model explainability tools were leveraged to determine which peaks were most significant for predicting HbA1c.

### CNN

A Convolutional Neural Network (CNN) was constructed based on an architecture adapted from McHardy et al. [28], utilizing PyTorch and PyTorch Lightning in a Python 3.11.1 environment. The model’s architecture featured two sequential units, each containing a one-dimensional convolutional layer with 64 filters and a kernel size of 5. This was followed by a rectified linear unit (ReLU) activation, batch normalization, and a max-pooling layer with a pool size of 2.

Data pre-processing for the CNN mirrored the procedure used for PLSR (Figure 2A). Initially, spectral regions of 800–1800 cm⁻¹ and 2800–3400 cm⁻¹ were selected for their biological relevance.

Following the convolutional units, the output was flattened and passed to two fully connected dense layers with ReLU activations and dropout rates of 0.1 and 0.2, respectively. The final output layer consisted of a single neuron with a linear activation function to predict HbA1c levels. The model was trained using the Adam optimizer with Mean Squared Error (MSE) as the loss function with a learning rate of 0.0001. To improve training stability, the learning rate was halved if the validation loss failed to improve for 10 consecutive epochs. Additionally, an early stopping criterion terminated the training process after 15 epochs without improvement, restoring the model weights from the best-performing epoch.

### Models quality evaluation

Several statistical metrics were employed to assess the accuracy and reliability of the models developed for HbA1c prediction using FTIR spectra. These metrics include the Pearson correlation coefficient (r), the root mean squared error (RMSE), mean absolute error (MAE), the mean squared error (MSE), the coefficient of determination (R²), and Discrepancy (DIS) (Supplementary Information).

### SHAP Values

Explainable artificial intelligence approaches, such as SHAP (SHapley Additive exPlanations), were employed to improve the interpretability of the H2O AutoML model used in this study. SHAP is a model-agnostic method grounded in cooperative game theory, assigning each feature an importance value for a particular prediction. In the context of spectral data, each wavelength (or spectral feature) is treated as a player contributing to the prediction outcome. SHAP values quantify how much each spectral variable contributes to increasing or decreasing the predicted output (e.g., HbA1c level), thus offering a transparent, feature-level explanation of model behavior. Importantly, while SHAP enhances interpretability by identifying influential features, it does not assess the predictive accuracy or quality of the model itself. The SHAP analysis was performed as follows. First, the marginal contribution of each feature was computed, assigning a numeric SHAP value to each feature for every prediction. A positive SHAP value indicates that the feature increased the predicted outcome, whereas a negative value suggests a decreasing effect. The expected value, or baseline prediction, was determined as the average model output across all samples.

To evaluate overall feature importance, SHAP values were then aggregated across the dataset by calculating the mean absolute SHAP value for each feature. This approach allowed identification of the wavelengths with the highest influence on the model’s predictions.

### Software

The work was conducted using Python version 3.11.4. Numerical computations used NumPy. The regression model was implemented using the scikit-learn module, while statistical analysis and signal smoothing were performed with the scipy module. Polynomial curve fitting was accomplished with the JAXFit module, which leverages GPU/TPU acceleration for computations [28]. Additionally, a regression model was developed using H2O AutoML. Each curve fitting procedure was executed on a Google Colab T4 GPU, with an average computation time of approximately 1.5 minutes per sample. SHAP and permutation feature importance analyses were performed using the built-in functionality of the H2O AutoML.

## Results

### Performance of HbA1c Prediction Models

The study included 673 patients samples which met the eligibility criteria (Figure 1). Three analytical approaches – partial least squares regression (PLSR), convolutional neural networks (CNNs), and curve fitting (CF) – were applied to analyze FTIR spectra of patients’ blood. As summarized in Table 3, the curve fitting model showed weaker predictive performance (R² = 0.59) compared to PLSR (R² = 0.76) and CNN (R² = 0.76).

**Table 2.** FTIR absorption peak assignments for blood samples from patients with and without diabetes.

| Bio-fingerprint region ($\approx 1000\text{--}1225\text{ cm}^{-1}$) | | | |
| --- | --- | --- | --- |
| Wavenumber ($\text{cm}^{-1}$) | Vibrational mode | Biochemical assignment | References |
| 930 | C–C skeletal vibrations | DNA (Z-form), nucleic acids, carbohydrates | [29] |
| 986 | C=C out-of-plane bending; C3N stretching (porphyrins), C–C DNA backbone | Lipids, alkenes, porphyrins, DNA | [30], [59] |
| 1011 | C–O–P stretch; C–O stretch + C–O bend (C–OH groups) | DNA phosphates, carbohydrates | [31] |
| 1023 | C–O stretching | Carbohydrates, glycoproteins, phosphate groups; C–C in phospholipids; glycogen | [32], [60] |
| 1030 | C–O / C–N stretching | Phosphates & carbohydrates; glycogen; protein glycation | [33] |
| 1051–1055 | C–O stretching; $\text{PO}_2^-$ symmetric / $\text{PO}_3^{2-}$ asymmetric bands | Phospholipids; nucleotides; lipids; cholesterol; glucose; riboflavin-5-phosphate | [34], [61] |
| 1080–1084 | hydroxyl groups, $\nu(\text{C–O})$, $\text{PO}_2^-$ symmetric stretch | glycogen/glucose/Phospholipids and proteins | [35] |
| 1104 | C–O stretch or $\text{CH}_2$ bend; $\nu(\text{C=O})$ of ribose/RNA | Carbohydrates/fatty acids; glycogen C–O + glucose O–H deformation | [36] |
| 1122 | C–C in lipids; N–H bend / C–N stretch in proteins | D-mannose; glucose; ribose ($\sim 1117$) C–O; heme C-methyl; carbohydrate carbonyls | [37], [62], [79] |
| 1165 | C–O–C stretching; C–C stretching | Carbohydrates/lipids; C–O from Ser/Thr/Tyr; hemoglobin ($\sim 1169$) | [38] |
| 1212 | C–H deformations; $\text{PO}_2^-$ asymmetric stretch | Oxy/deoxy-hemoglobin; nucleic acids; phospholipids | [39] |
| 1223 | $\text{PO}_2^-$ asymmetric stretch; DNA conformational marker | B-form DNA; oxyhemoglobin; nucleic acids | [40], [63] |
| Amide III & nearby ($\approx 1230\text{--}1350\text{ cm}^{-1}$) | | | |
| Wavenumber ($\text{cm}^{-1}$) | Vibrational mode | Biochemical assignment | |
| 1239 | Amide III: C–N stretch + N–H bend; $\text{CH}_2$ wag | Proteins; may overlap with $\text{PO}_2^-$ (sym/asym) | [41], [64] |
| 1302–1304 | Amide III: N–H bend + C–N stretch; CH deformation (Leu) | Proteins; carbohydrates/glucose contributions; hemoglobin context | [42], [65], [80] |
| 1313 | Amide III: $\text{C=O}$; C–H deformation | Protein backbone/secondary structure | [43] |
| CH bending region ($\approx 1350\text{--}1460\text{ cm}^{-1}$) | | | |
| Wavenumber ($\text{cm}^{-1}$) | Vibrational mode | Biochemical assignment | |
| 1366 | $\text{CH}_3$ symmetric deformation; $\text{CH}_2$ bending | Lipids; serum proteins; glycoproteins | [44], [66] |
| 1390–1394 | $\text{CH}_3$ Symmetric stretching; $\text{COO}^-$ symmetric stretch | Fatty acids | [45], [67], [81] |
| 1402 | $\text{CH}_3$ symmetric bending; $\text{COO}^-$ symmetric stretch/deformation | Protein methyl motions; carboxylates | [46], [68] |
| 1452 | In-plane C–H bending; $\text{CH}_2$ scissoring | Lipids/fatty acids; protein C–H bending | [47], [69], [82] |

| Amide II ($\approx 1500\text{--}1580\text{ cm}^{-1}$) | | | |
| --- | --- | --- | --- |
| Wavenumber ($\text{cm}^{-1}$) | Vibrational mode | Biochemical assignment | |
| 1539 | Amide II: C–N stretch + N–H bend (CN/CNH bend; N–H stretch noted) | Proteins; Trp bands; Hb states; metabolic stress markers context | [48], [70], [83] |
| 1560 | Amide II: in-plane N–H bend + C–N stretch; possible $\text{CO}_2^-$ asym (Glu) | IgG; collagen I; general proteins | [49], [71], [84], [87] |
| 1574 | vas($\text{COO}^-$) (Asp/Glu); possible aromatic C=C | Amino-acid side chains (Asp); nucleic bases contribution | [50], [72] |
| Amide I ($\approx 1600\text{--}1700\text{ cm}^{-1}$) | | | |
| Wavenumber ($\text{cm}^{-1}$) | Vibrational mode | Biochemical assignment | |
| 1641 | Amide I: C=O stretching (water near 1646) | Protein secondary structure; hydrated water contribution | [51], [73], [85], [88] |
| 1660 | Amide I sub-band | α-helix / random coil (context-dependent); collagen/albumin/globulins | [52], [74] |
| 1678 | Amide I sub-band | β-turns; also C=O ($\text{CONH}_2$) in Asn side chains | [53], [75] |
| 1687 | Amide I high-wavenumber component | β-turns / antiparallel β-sheets / aggregates (context-dependent) | [54], [76], [86] |
| CH stretching region ($\approx 2800\text{--}3000\text{ cm}^{-1}$) | | | |
| Wavenumber ($\text{cm}^{-1}$) | Vibrational mode | Biochemical assignment | |
| 2872 | vs($\text{CH}_3$) | Lipids; (also present in proteins) | [55] |
| 2930 | vas($\text{CH}_2$) | Lipids; fatty acids; cholesterol region | [56] |
| 2953 | vas($\text{CH}_3$) | Lipids; lipid-rich environments | [57] |
| Amide A / O–H ($\approx 3294\text{ cm}^{-1}$) | | | |
| Wavenumber ($\text{cm}^{-1}$) | Vibrational mode | Biochemical assignment | |
| 3294 | N–H stretching (Amide A); O–H stretching (H-bonded) | Proteins; hydration/water contribution | [58], [78], [150] |

**Table 3.**
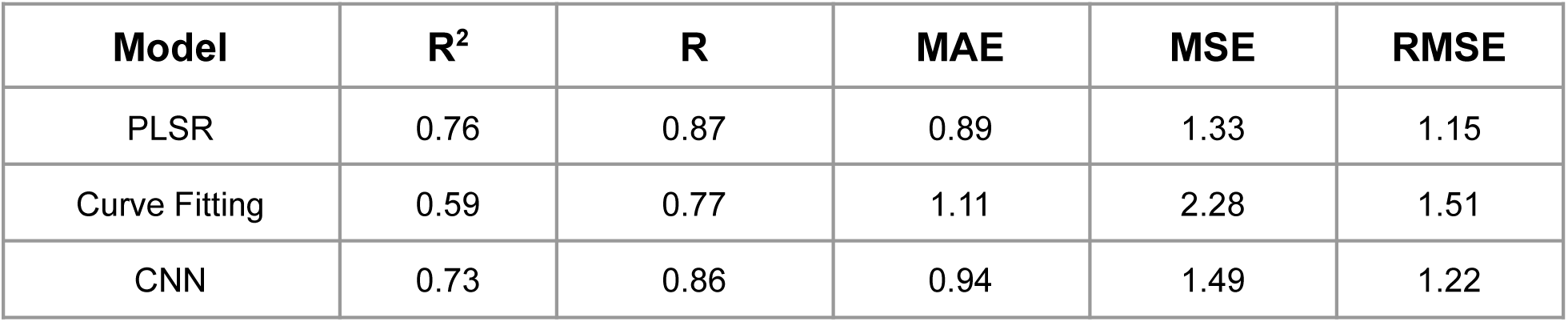
Model performance on test data comparison for PLSR, CF, and CNN.

The PLSR model was performed on the 800–1800 cm⁻¹ and 2800–3400 cm⁻¹ regions and optimized using 10-fold cross-validation. Model complexity was selected based on the Q² criterion, where Q² denotes the cross-validated predictive ability of the model. This procedure identified nine latent components as optimal. Applied to the independent test set, this configuration achieved an R² of 0.76, RMSE = 1.16, and MAE = 0.89 (Figure 4).

**Figure 4.**
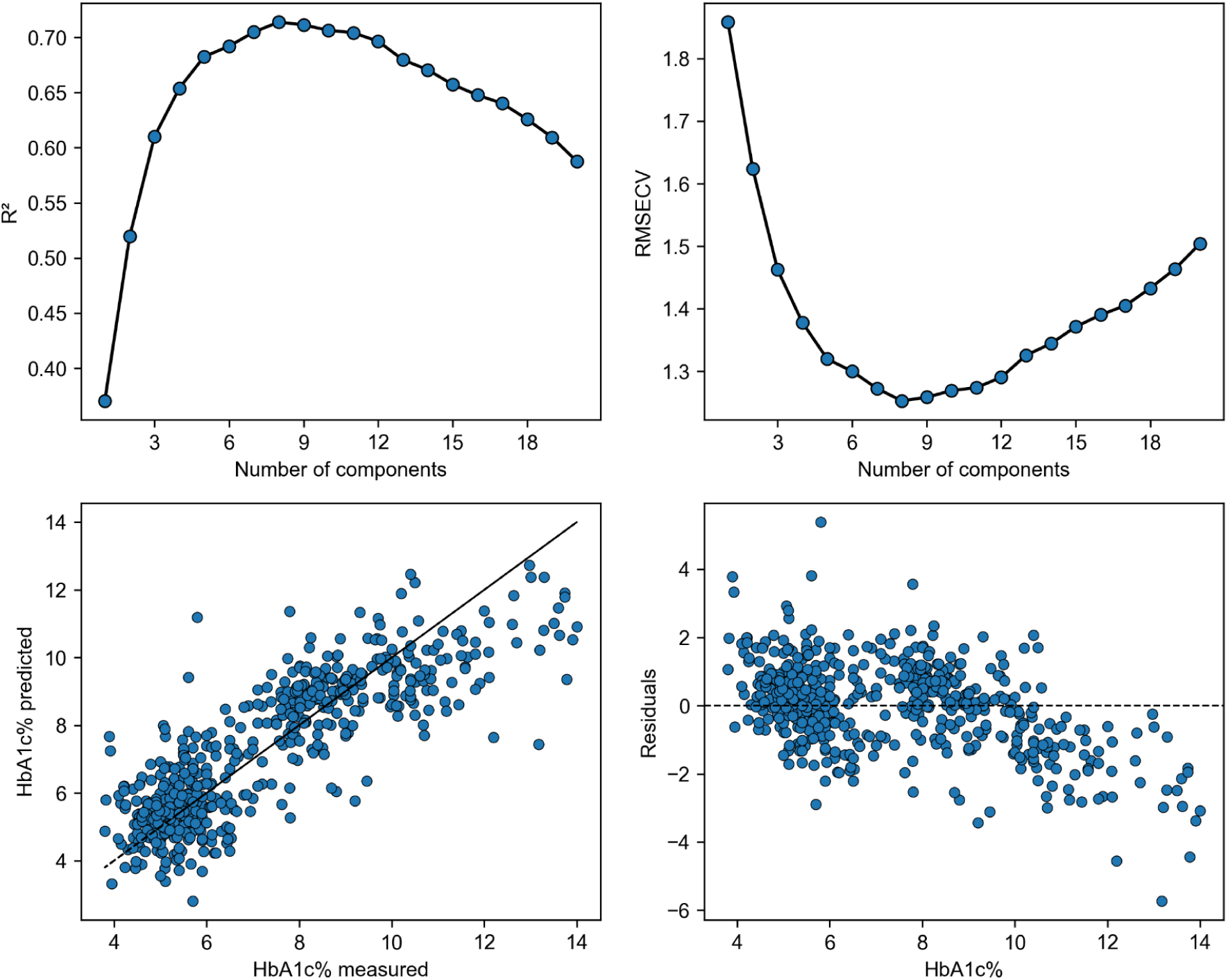
PLSR model. Results of the training calibration data (CV10). The top row displays the dependency of the coefficient of determination (R²) and RMSECV (Root Mean Square Error of Cross-Validation) on the number of components. The bottom row presents the graph of measured HbA1c versus predicted values (left) and the residuals after prediction (right).

Curve fitting models were developed from parameters extracted within the 1000–1720 cm⁻¹ region, providing high-quality fits to individual spectra (R² > 0.9999). Among the evaluated parameters, peak height offered the best performance; however, the final model remained less predictive than PLSR and CNN, with R² = 0.59, RMSE = 1.51, and MAE = 1.11 on the test set (Figure 5B).

**Figure 5.**
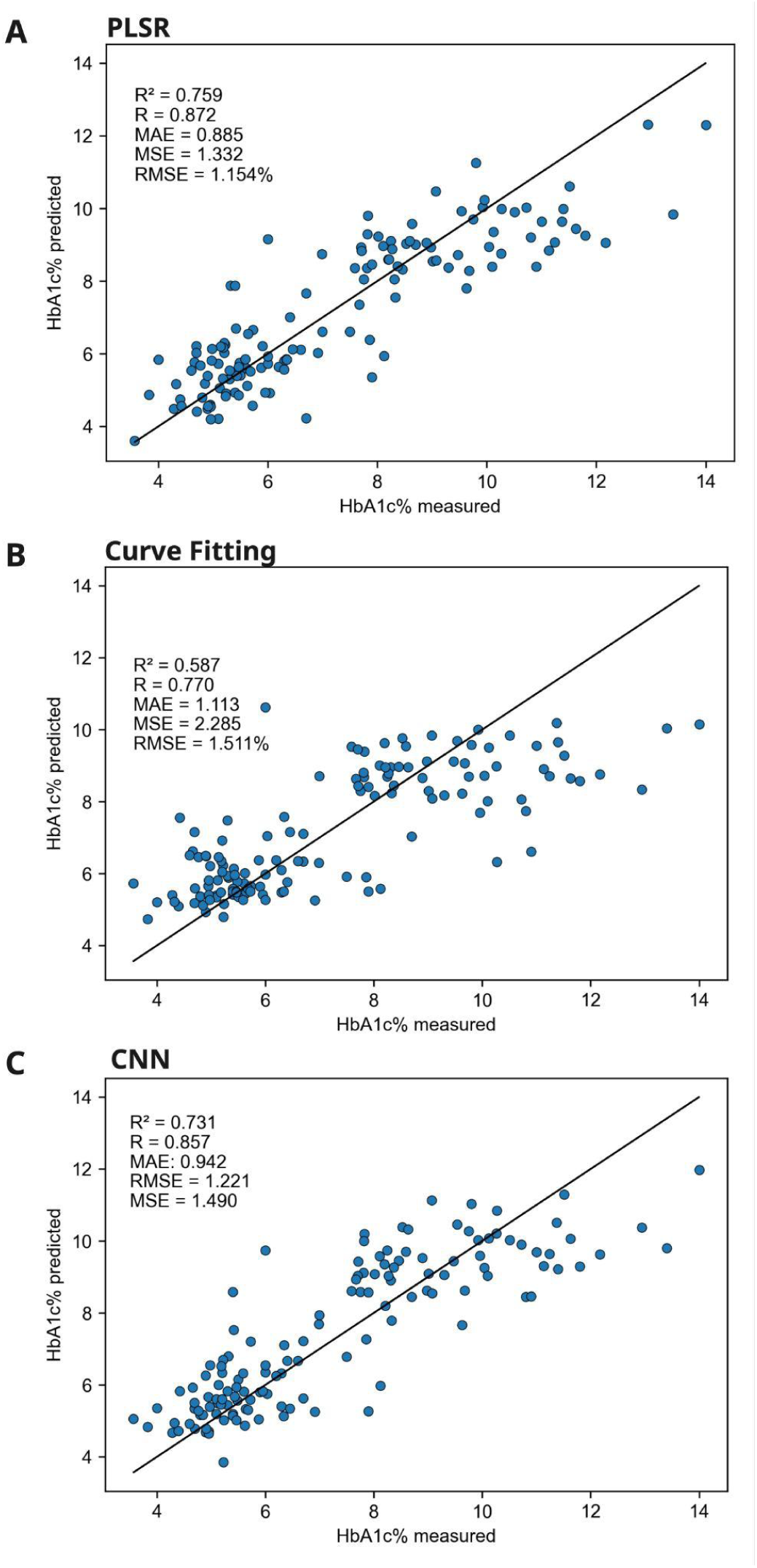
Models perfomance: (A) PLSR performance on the test data (B) Validation plot of the best H2O AutoML model, showing predicted HbA1c% against measured HbA1c%. The proximity of data points to the main diagonal line reflects prediction accuracy, supported by the displayed error metrics. (C) CNN validation plot HbA1c measured vs HbA1c predicted by CNN model.

The CNN model, trained on the same spectral regions as PLSR, achieved an R² of 0.73, RMSE = 1.22, and MAE = 0.94 indicating that a substantial amount of HbA1c-related spectral information can be learned without explicit peak engineering (Figure 5C).

Because CNN did not substantially outperform PLSR, the key predictive patterns in the selected FTIR regions appear to be largely linear, while nonlinear effects may play a secondary role under the present experimental conditions. Region-wise attribution revealed that CNN predictions relied primarily on the fingerprint carbohydrate region (∼1000–1100 cm⁻¹), the amide I–II bands (∼1500–1700 cm⁻¹), and CH-stretching (∼2800–3000 cm⁻¹), consistent with glycation-related biochemical changes highlighted by curve fitting and SHAP.

A direct comparison of the three modeling strategies (Table 3) shows that while PLSR achieved the highest predictive accuracy, CNN performed comparably and consistently outperformed curve fitting. These results emphasize the balance between interpretability and predictive strength: curve fitting offers clear biochemical meaning at the peak level, whereas PLSR and CNN capture broader or nonlinear spectral relationships that improve generalization.

### Identification of Spectral Features Influencing HbA1c Prediction

Next, we were aiming to identify the spectral features that most strongly influenced the prediction, the entire spectral range was examined in reference to literature data to determine the available peaks and their biochemical assignments. Table 2 summarizes the major FTIR absorption peak assignments observed in the study. The spectra revealed characteristic bands distributed across the bio-fingerprint region (≈1000–1225 cm⁻¹), amide III region (≈1230–1350 cm⁻¹), CH-bending region (≈1350–1460 cm⁻¹), amide II region (≈1500–1580 cm⁻¹), amide I region (≈1600–1700 cm⁻¹), CH-stretching region (≈2800–3000 cm⁻¹), and the amide A/O–H region (≈3294 cm⁻¹).

To detect the most influential peaks, fits and final models were obtained with Curve fitting as described in Materials and Methods. The method allowed to yield peak parameters (position, normalized area, FWHM, η, height) for HbA1c prediction. An example fit is shown in Figure 6.

**Figure 6.**
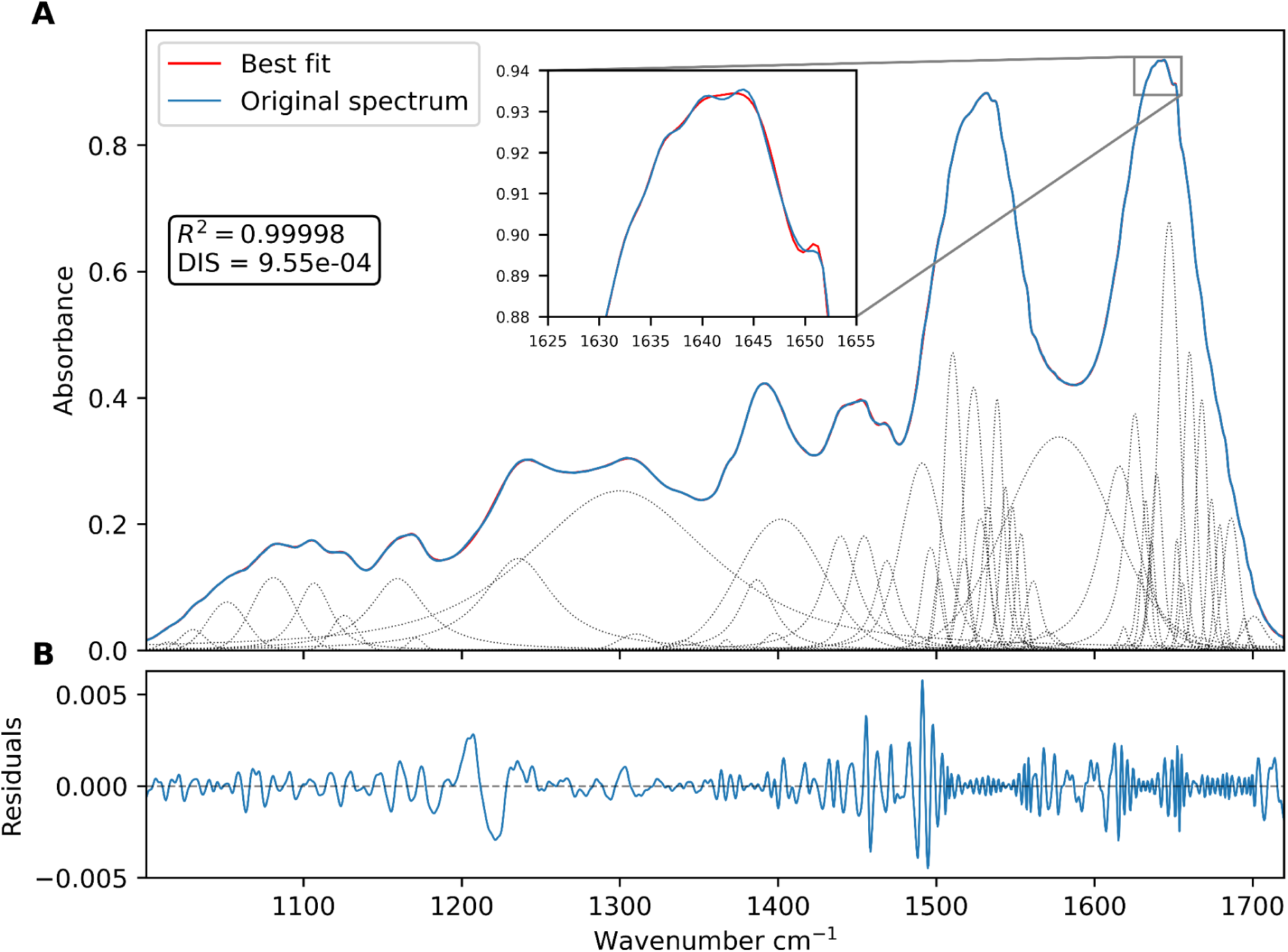
Curve fitting of the blood FTIR spectrum. (A) the spectrum in the 1000–1700 cm⁻¹ region. The original experimental spectrum (blue line) is deconvoluted into multiple Gaussian/Lorentzian components (grey dotted lines), which together reconstruct the best-fit spectrum (red line). The high quality of the fit is demonstrated by the coefficient of determination (R² = 0.99999) and the low discrepancy (DIS = 8.88 × 10⁻⁴). The zoom in highlights the amide I region (1625–1655 cm⁻¹), showing the close overlap between the experimental and fitted curves, confirming the accuracy of the deconvolution. (B) The residual plot demonstrates consistently low values, remaining within ±0.004 a.u. across the entire spectrum. While minor structural deviations exist in regions of high peak density, the low amplitude confirms the deconvolution quality.

Variable importance was subsequently extracted from the top 20 AutoML models. Figure 7 illustrates the distribution of the most influential peaks, ranked by median feature importance. The importance values displayed here come from each model’s internal metrics and indicate how strongly individual peaks contributed to overall prediction accuracy. Although model-agnostic feature ranking methods such as permutation importance can estimate how prediction error changes when features are randomly shuffled, they do not convey whether a given feature increases or decreases predicted HbA1c levels. Because directionality was essential to our analysis, we applied SHAP (Shapley additive explanations) values, which provide both the magnitude and the direction of each feature’s contribution to the model output.

**Figure 7.**
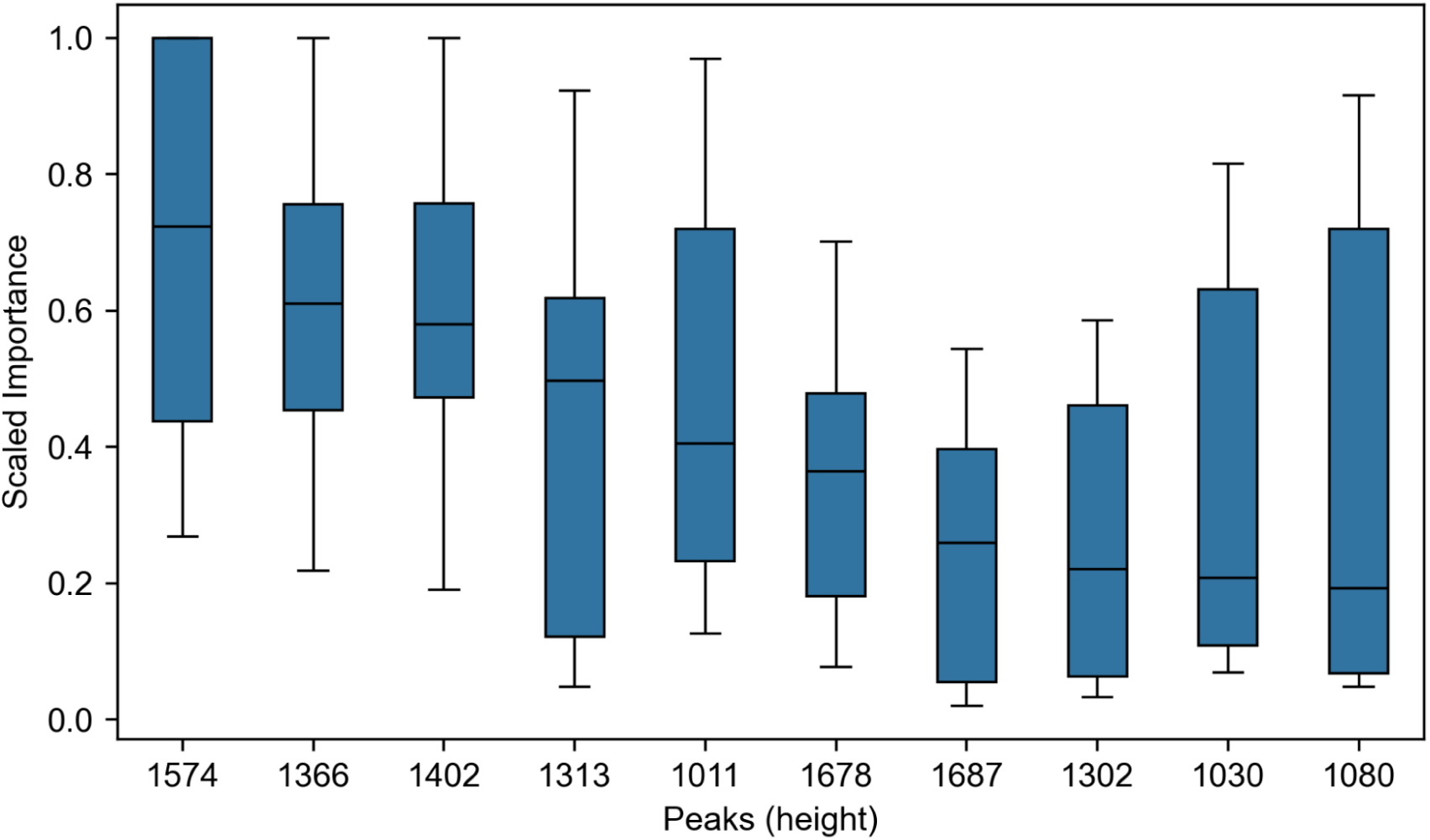
Box plot displaying the scaled importance of the top 10 identified peaks from the 20 best-performing H2O AutoML models.

The interpretation of the peaks featured with SHAP is presented in Table X. The first two columns rank the top 10 spectral peaks identified by the 20 best-performing H2O AutoML models and the 20 most influential features derived from SHAP value analysis of the best model. The subsequent columns list the wavenumber positions (cm⁻¹), their vibrational assignments, and the biochemical interpretations based on SHAP-derived pattern relationships.

SHAP captures both the magnitude and direction of feature contributions at local (sample-level) and global scales (Figure 8). Using this approach, we examined the peaks prioritized by curve fitting, revealing clear biochemical patterns: lipid- and protein-related vibrations (e.g., 1574, 1402, 1366 cm⁻¹), carbohydrate- and glycation-specific regions (1011, 1030, 1230 cm⁻¹), and secondary-structure–sensitive amide bands (1678–1687 cm⁻¹) all showed distinct associations with HbA1c levels.

**Figure 8.**
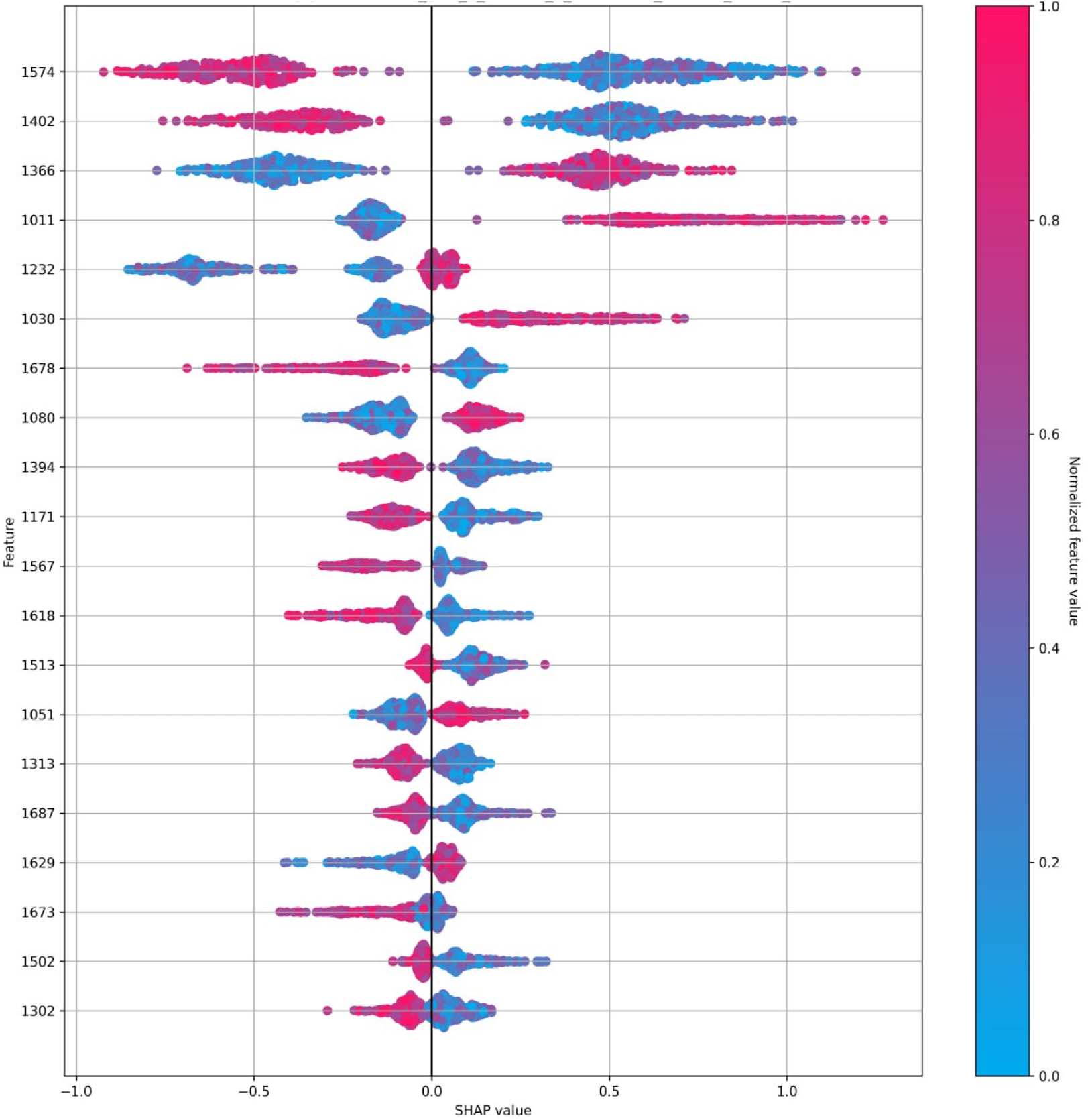
Beeswarm plot displaying the 20 most influential features based on SHAP value analysis from the best H2O AutoML model. This visualization highlights the distribution of SHAP values for each top-ranked feature across all samples, illustrating both the magnitude and direction of their impact on model predictions. The plot provides an intuitive overview of how individual features contribute to variability in the predicted outcomes.

## Discussion

The infrared spectrum of blood exhibits a complex fingerprint region that reflects the major biomolecular components: proteins, lipids, carbohydrates, and nucleic acids. In our study the typical spectra were observed and analysed. Namely, 1000–1225 cm⁻¹ region, bands primarily arise from phosphate–sugar vibrations, including C–O and C–C stretching modes of carbohydrates, glycogen, and nucleic acids [89, 90]. The amide III region (1230–1350 cm⁻¹) reflects combined C–N stretching and N–H bending, providing insight into protein secondary structures [91]. The CH bending region (1350–1460 cm⁻¹) originates from CH₂ and CH₃ deformations, with contributions from both proteins and lipids [92]. In the amide II (1500–1580 cm⁻¹) and amide I (1600–1700 cm⁻¹) regions, characteristic vibrations of the peptide backbone dominate: N–H bending/C–N stretching and C=O stretching, respectively, allowing evaluation of α-helix, β-sheet, and random coil structures [93]. Finally, the CH stretching region (2800–3000 cm⁻¹) reports on lipids and fatty acids, while the amide A/OH region (∼3290 cm⁻¹) corresponds to N–H and O–H stretching, influenced by protein conformation and hydrogen bonding [94].

Aiming to enhance HbA1c diagnostic performance, we focused on FTIR-derived features associated with hemoglobin structure, glycation-related sugar moieties, and protein modification patterns in blood samples. Therefore, among the identified peaks, the following were considered hemoglobin-associated: 1539, 1560, 1452, 1390–1394, 1169, 1313, 1088, and 932 cm⁻¹. Amide II bands were observed at 1539 and 1560 cm⁻¹, corresponding to C–N stretching and N–H bending vibrations of the hemoglobin backbone [95, 69, 81]. In the mid-fingerprint region, the band at 1452 cm⁻¹, assigned to CH₂ bending, reflects contributions from protein side chains and has been linked to structural changes of hemoglobin in type 2 diabetes. The 1390–1394 cm⁻¹ band is attributed to CH₃ symmetric deformation, mainly arising from carboxylate groups formed by acidic amino acids such as glutamic acid and aspartic acid. The band at 1165 cm⁻¹, assigned to C–O–C and C–C stretching, reflects contributions from carbohydrates, lipids, and residues such as serine, threonine, and tyrosine, as well as hemoglobin itself (∼1169 cm⁻¹). Aromatic amino acids, including tryptophan and tyrosine, contribute to the signal near 1313 cm⁻¹, which corresponds to C–N stretching vibrations of aromatic groups in the peptide chain. A band at 1088 cm⁻¹ represents C–O stretching associated with carbohydrates, phospholipids, and nucleic acid backbones. Finally, a feature near 932 cm⁻¹ was assigned to C–C–(N) stretching, typically linked to DNA backbone vibrations, although partial overlap with hemoglobin cannot be excluded [96, 97].

Several peaks associated with glucose and carbohydrate structures were also identified. The band at around 930 cm⁻¹ corresponds to C–C–(N) stretching and is commonly linked to carbohydrate backbone vibrations. A broader region between 986 and 1055 cm⁻¹ reflects C–O stretching in C–OH groups and C–C stretching within carbohydrate structures, consistent with signals from glucose and related saccharides. In addition, the band at 1104 cm⁻¹ is assigned to C–O stretching in C–O–C linkages, characteristic of glycosidic bonds in carbohydrates [98].

The absorption band at approximately 1030 cm⁻¹ arises from C–O and C–N stretching vibrations. In studies quantifying protein glycation using vibrational spectroscopy, this peak was observed exclusively in glycated samples, where it was attributed to covalently bound glucose residues. Importantly, the intensity of the 1030 cm⁻¹ band was shown to correlate directly with the degree of glycation, highlighting its value as a spectral marker of glucose–protein adduct formation [99].

It was found that increase in absorbance at 1396 cm⁻¹ reflects enhanced vibrations of the carboxylate (COO⁻) groups of aspartic acid residues. Among these, Aspβ99 plays a particularly critical role as part of the α1β2 subunit interface, a region that governs the cooperative transition of hemoglobin between its low- and high-affinity states. In this “switch” region, Aspβ99 forms a stabilizing hydrogen bond with Tyrα42, and modulation of this interaction is essential for the structural rearrangements underlying allosteric regulation. Thus, the spectral feature at 1396 cm⁻¹ provides a molecular readout of conformational dynamics at one of hemoglobin’s most functionally important interfaces [100, 101].

Although previous studies have associated bands near 1710 cm⁻¹ (carbonyl stretches) and ∼1620 cm⁻¹ (amide-I sub-band often linked to aggregated/β-rich structures) with the gradual formation and deposition of irreversible AGE products, these features were not prominent in our patient blood spectra. We attribute this to the low fractional abundance. In addition, our cohort’s HbA1c distribution likely reflects a predominance of early glycation (ketoamines) rather than extensive crosslinking [102].

Table 3 summarizes the key FTIR spectral bands identified as relevant to HbA1c prediction, based on SHAP and describes possible biochemical alterations related to early glycation chemistry to structural modifications of proteins and lipids.

**Table 3.**
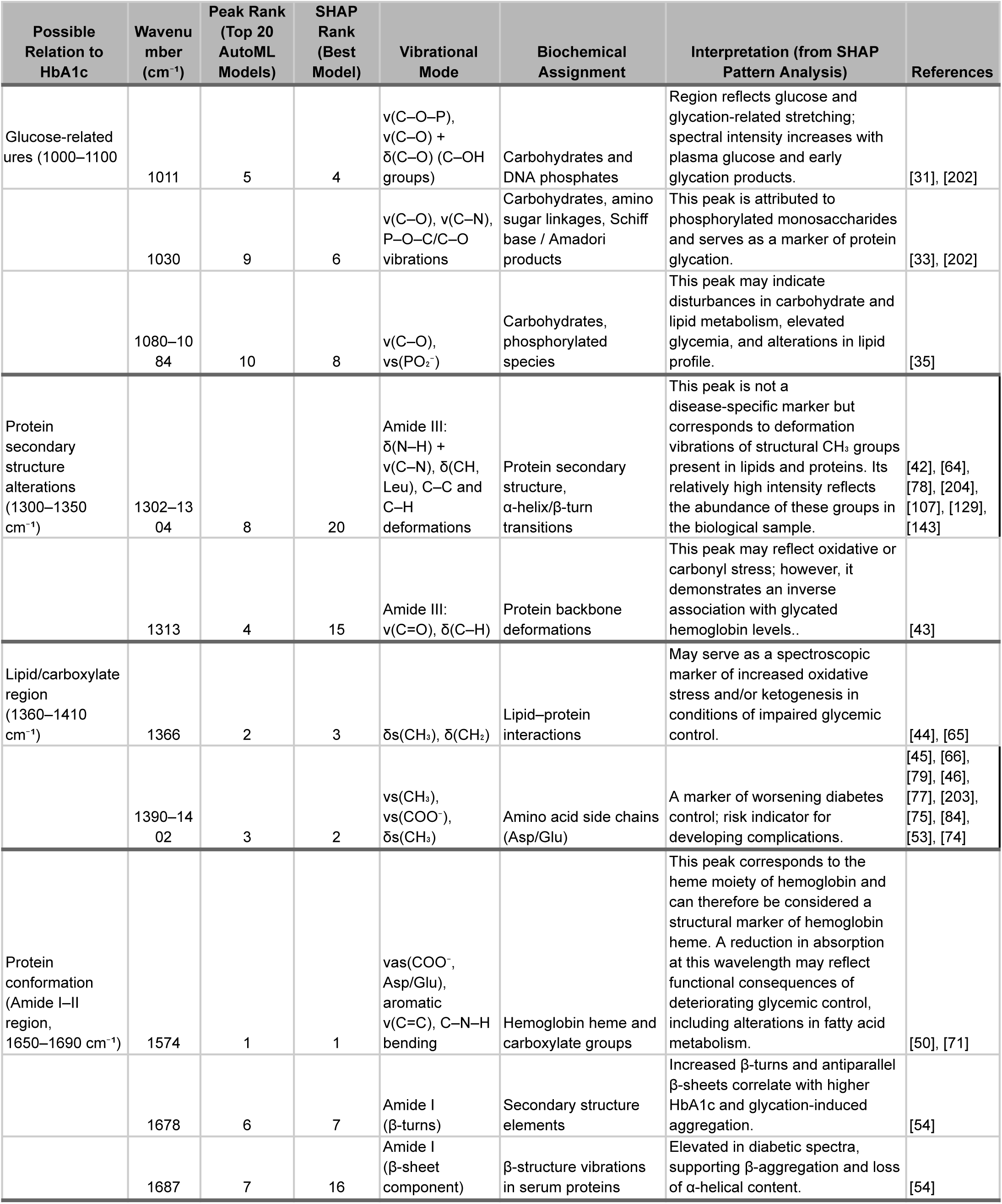
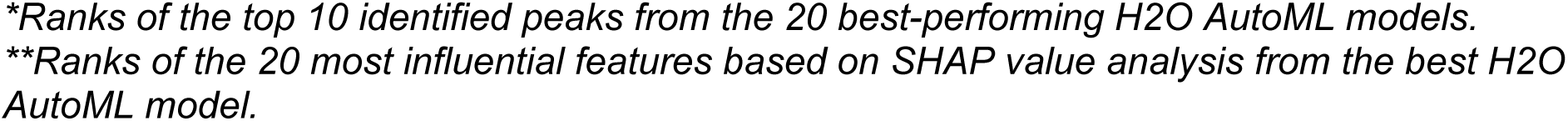
FTIR spectral bands identified as relevant to HbA1c prediction.

Bands within the 1000–1100 cm⁻¹ region (1011, 1030, 1080 cm⁻¹) correspond to C–O and C–N stretching vibrations of carbohydrates and phosphate groups, characteristic of early glycation and glucose-derived adducts. Their enhanced absorbance with rising HbA1c reflects increasing levels of covalently bound glucose residues.

The Amide III region (1300–1350 cm⁻¹) exhibits strong HbA1c dependence, particularly at 1302 cm⁻¹ and 1313 cm⁻¹, which mark secondary-structure rearrangements in hemoglobin and serum proteins. Shifts and intensity changes here indicate loss of α-helical content and accumulation of β-turn and β-sheet motifs associated with protein glycation.

Within the CH bending and Amide I–II regions (1360–1690 cm⁻¹), peaks at 1366, 1390–1402, 1574, 1678, and 1687 cm⁻¹ capture lipid–protein interactions and conformational remodeling. The 1366 cm⁻¹ band arises from CH₂/CH₃ deformations, suggesting altered lipid composition.

### Clinical and Diagnostic Relevance

FTIR combined with advanced analytical tools allows interpreting the results. This study proves that the single regression approach is lacking information, whereas the combinatory approach holds the promise for accurate diagnostic and prognostic data in the realm of clinical biochemistry.

Compared with conventional HPLC, which is optimized to deliver highly specific but typically single-analyte measurements such as HbA1c, FTIR spectroscopy provides a comprehensive biochemical fingerprint from the same blood sample. Each HPLC assay generally requires different columns, gradients, and reference standards, meaning that separate runs are often necessary to obtain additional parameters such as lipid levels or protein modifications. In contrast, a single FTIR spectrum encodes information across multiple molecular classes simultaneously, including glycated proteins, lipids and fatty acids, protein secondary structures, and nucleic acids. This multiplexing capacity allows FTIR to deliver a broader biochemical profile from one measurement, highlighting its potential not only as an HbA1c surrogate but also as a versatile screening platform capable of revealing additional clinically relevant markers in a cost- and time-efficient manner [103, 104, 105].

Despite promising results, several methodological limitations of FTIR-based modeling should be considered. By itself, FTIR spectroscopy has inherent constraints: strong water absorption can obscure parts of the spectrum, baseline variations may arise from sample thickness and scattering, and overlapping vibrational bands often complicate straightforward assignments. While preprocessing and deconvolution improve resolution, residual ambiguities in band attribution—such as distinguishing glycation-specific peaks from overlapping lipid or nucleic acid contributions—remain a challenge [106–108].

With respect to modeling approaches, PLSR provides robustness and interpretability but assumes linear relationships, which may not fully capture the complexity of biochemical interactions [109, 110]. Curve fitting offers high interpretability at the peak level, yet its reliance on assumptions about peak shape (e.g., pseudo-Voigt functions) and arbitrary initialization can limit accuracy and reproducibility, particularly in noisy spectra [111, 112]. CNNs overcome linearity constraints and capture multi-scale spectral features, but they are computationally intensive and less transparent, with interpretability hinging on post hoc explainability methods [113–116]. While Layer-wise Relevance Propagation (LRP)[117] and similar techniques can provide local feature attribution for CNNs, their outputs often correspond to spectral regions that remain superpositions of multiple vibrational modes, limiting biochemical specificity.

In addition, this study was limited by sample size and cohort heterogeneity, which may restrict generalizability. Larger and more diverse datasets are needed to confirm the robustness of the models across populations and clinical conditions. Peak fitting also remains non-ideal: convergence depends on initialization and parameter constraints, and in regions with overlapping peaks the deconvolution may not reflect a unique physical reality.

### Complementary approach benefits

The application of multiple machine learning approaches provided complementary perspectives on the biochemical information contained in FTIR spectra. Each method captured distinct yet converging aspects of HbA1c-related variability: curve fitting offered direct molecular interpretability by identifying specific glycation-sensitive peaks; PLSR extracted linear latent structures that summarized correlated spectral changes across broader biochemical regions; and CNN learned nonlinear and higher-order spectral patterns beyond classical regression. These findings suggest that integrating these approaches could enable a more comprehensive interpretation of glycation-associated spectral changes by linking interpretable molecular vibrations with statistical generalization. This concept aligns with multi-model strategies explored in other spectroscopic applications. For Raman spectra of chemical mixtures, comparisons between PLS and 1D-CNN models showed that both approaches achieved comparable accuracy, with PLS performing best on small, low-noise datasets and CNNs excelling in larger, noisier regimes [118]. In mid-infrared glucose analysis, combining Lorentzian profile-based curve fitting with support vector regression demonstrated that peak-resolved features derived from physically meaningful fits can substantially improve machine-learning performance [119]. Similarly, other researchers have developed a multi-model estimation framework in which FTIR-based predictions are obtained by averaging several chemometric and neural models, yielding higher accuracy and robustness than any single model [120]. Consequently, adopting such a strategy for HbA1c monitoring could enhance clinical translation by combining the mechanistic insight from curve fitting with the predictive strength of PLSR and deep learning, ultimately improving biomarker reliability and physiological interpretability.

Building on these findings, future work may explore hybrid approaches that combine the strengths of different methods – for example, using curve fitting to generate biochemically meaningful features that are subsequently modeled with PLSR or CNNs, or embedding physically constrained priors into deep-learning architectures. Advances in explainable AI, including SHAP, LRP, and Bayesian frameworks, may further enhance the interpretability and reliability of predictions [116, 121]. Ultimately, addressing these methodological and sample-related limitations will strengthen the clinical utility of FTIR spectroscopy for HbA1c monitoring and expand its applicability to other biomarkers.

## Conclusion

This study demonstrates the efficacy of Fourier-transform infrared (FTIR) spectroscopy combined with machine learning for the rapid assessment of HbA1c. While Partial Least Squares Regression (PLSR) and Convolutional Neural Networks (CNN) achieved the highest predictive accuracy (R² = 0.76 and 0.73, respectively), Curve Fitting (CF) provided essential mechanistic validation. Despite lower predictive power (R² = 0.59), CF successfully deconvoluted complex spectra to identify specific glycation-sensitive features, including lipid and protein vibrations (1574, 1402, 1366 cm⁻¹) and carbohydrate bands (1011 cm⁻¹) confirmed by SHAP analysis.

The results indicate that these modeling strategies are not mutually exclusive but complementary: PLSR and CNN capture broad linear and nonlinear spectral patterns necessary for generalization, whereas peak-resolved analysis ensures biochemical interpretability. This multi-model framework supports FTIR spectroscopy as a scalable, reagent-free alternative to HPLC for large-scale diabetes screening. Future implementation should focus on hybrid architectures that integrate physically constrained features into deep learning models to further enhance diagnostic reliability and clinical translation.

## Author Contributions

MM and TM contributed equally to this work. MM conducted the computational experiments, prepared the datasets, and developed the analysis scripts. BD performed data acquisition. DK and TM coordinated the study, critically evaluated the results, and supervised the analysis. TM performed sample processing and spectral quality assessment, while KM performed peak characterization. All authors contributed to the drafting of the manuscript.

## Data Availability

The patient-level data, including raw FTIR spectra and associated clinical measurements, are not publicly available due to ethical and privacy restrictions established by the Ethics Committee of the State Institution “D.F. Chebotarev Institute of Gerontology of the National Academy of Medical Sciences of Ukraine.” De-identified data may be made available from the corresponding author upon reasonable request and subject to institutional approval. The source code for spectral preprocessing and model development is openly available at https://github.com/MelnychenkoM/ftir-hba1c-prediction.

## Ethical Statement

The study protocol No. 7 dated 11/07/2024 was approved by the Ethics Committee of the State Institution “D.F. Chebotarev Institute of Gerontology of the National Academy of Medical Sciences of Ukraine.” Written informed consent was obtained from each participant before the study began. All individuals involved, including family physicians, participated in the study as volunteers.

## References

1. Koenig, R. J. et al. Correlation of glucose regulation and hemoglobin A1c in diabetes mellitus. N. Engl. J. Med. 295, 417–420 (1976).

2. Goldstein, D. E. et al. Clinical application of glycosylated hemoglobin measurements. Diabetes 31 (Suppl. 3), 70–78 (1982).

3. Gabbay, K. H. et al. Glycosylated hemoglobins and long-term blood glucose control in diabetes mellitus. J. Clin. Endocrinol. Metab. 44, 859–864 (1977).

4. Larsen, M. L., Hørder, M. & Mogensen, E. F. Effect of long-term monitoring of glycosylated hemoglobin levels in insulin-dependent diabetes mellitus. N. Engl. J. Med. 323, 1021–1025 (1990).

5. Global Burden of Disease Collaborative Network. Global Burden of Disease Study 2021 Results. Institute for Health Metrics and Evaluation (IHME) (2024).

6. Ong, K. L. et al. Global, regional, and national burden of diabetes from 1990 to 2021, with projections of prevalence to 2050: a systematic analysis for the Global Burden of Disease Study 2021. Lancet 402, 203–234 (2023).

7. Ahmad, R. et al. Comparison of glycated hemoglobin (HbA1c%) between high performance liquid chromatography (HPLC) and non-HPLC methodology. J. Health Rehabil. Res. 4, 557–561 (2024).

8. Ye, S., Ruan, P., Yong, J. et al. The impact of the HbA1c level of type 2 diabetics on the structure of haemoglobin. Sci. Rep. 6, 33352 (2016).

9. Polakovs, M., Mironova-Ulmane, N., Kurjane, N., Reinholds, E. & Grube, M. Micro-Raman scattering and infrared spectra of hemoglobin. Proc. SPIE Int. Soc. Opt. Eng. 7142, 10.1117/12.815796 (2008).

10. Yin, H. et al. Wavenumber selection for FTIR/ATR spectroscopy analysis of hemoglobin in human whole blood. Proc. Int. Conf. Bioinform. Biomed. Eng. (ICBBE) 4, 1–4 (2010).

11. Sherwood, P. M. A. The use and misuse of curve fitting in the analysis of core X-ray photoelectron spectroscopic data. Surf. Interface Anal. 51, 589–610 (2019).

12. Sadat, A. & Joye, I. J. Peak fitting applied to Fourier transform infrared and Raman spectroscopic analysis of proteins. Appl. Sci. 10, 5918 (2020).

13. Jiang, S. et al. Using ATR-FTIR spectra and convolutional neural networks for characterizing mixed plastic waste. Comput. Chem. Eng. 155, 107547 (2021).

14. Ng, W. et al. Convolutional neural network for simultaneous prediction of several soil properties using visible/near-infrared, mid-infrared, and combined spectra. Geoderma 352, 251–267 (2019).

15. Acquarelli, J., van Laarhoven, T., Gerretzen, J., Tran, T. N., Buydens, L. M. C. & Marchiori, E. Convolutional neural networks for vibrational spectroscopic data analysis. Anal. Chim. Acta 954, 22–31 (2017).

16. Sánchez-Brito, M., Luna-Rosas, F. J., Mendoza-González, R., Mata-Miranda, M. M., Martínez-Romo, J. C. & Vázquez-Zapién, G. J. A machine-learning strategy to evaluate the use of FTIR spectra of saliva for a good control of type 2 diabetes. Talanta 221, 121650 (2021).

17. Enders, A. A., North, N. M., Fensore, C. M., Velez-Alvarez, J. & Allen, H. C. Functional group identification for FTIR spectra using image-based machine learning models. Anal. Chem. 93, 9711–9718 (2021).

18. Zhang, F. & Huang, Q. Characterization of Z-DNA by infrared spectroscopy. Methods Mol. Biol. 2651, 53–58 (2023).

19. Vázquez-Zapién, G. J., Mata-Miranda, M. M., Sánchez-Monroy, V., Delgado-Macuil, R. J., Pérez-Ishiwara, D. G. & Rojas-López, M. FTIR spectroscopic and molecular analysis during differentiation of pluripotent stem cells to pancreatic cells. Stem Cells Int. 2016, 6709714 (2016).

20. Movasaghi, Z., Rehman, S. & Rehman, I. U. Fourier transform infrared (FTIR) spectroscopy of biological tissues. Appl. Spectrosc. Rev. 43, 134–179 (2008).

21. Lesh, F. H. Multi-dimensional least-squares polynomial curve fitting. Commun. ACM 2, 29–30 (1959).

22. Sherwood, P. M. A. The use and misuse of curve fitting in the analysis of core X-ray photoelectron spectroscopic data. Surf. Interface Anal. 51, 589–610 (2019).

23. Sadat, A. & Joye, I. J. Peak fitting applied to Fourier transform infrared and Raman spectroscopic analysis of proteins. Appl. Sci. 10, 5918 (2020).

24. Jiang, S. et al. Using ATR-FTIR spectra and convolutional neural networks for characterizing mixed plastic waste. Comput. Chem. Eng. 155, 107547 (2021).

25. Ng, W. et al. Convolutional neural network for simultaneous prediction of several soil properties using visible/near-infrared, mid-infrared, and combined spectra. Geoderma 352, 251–267 (2019).

26. Acquarelli, J., van Laarhoven, T., Gerretzen, J., Tran, T. N., Buydens, L. M. C. & Marchiori, E. Convolutional neural networks for vibrational spectroscopic data analysis. Anal. Chim. Acta 954, 22–31 (2017).

27. Sánchez-Brito, M., Luna-Rosas, F. J., Mendoza-González, R., Mata-Miranda, M. M., Martínez-Romo, J. C. & Vázquez-Zapién, G. J. A machine-learning strategy to evaluate the use of FTIR spectra of saliva for a good control of type 2 diabetes. Talanta 221, 121650 (2021).

28. Hofer, L. R., Krstajić, M. & Smith, R. P. JAXFit: trust region method for nonlinear least-squares curve fitting on the GPU. arXiv preprint arXiv:2208.12187 [cs.LG] (2022). 10.48550/arXiv.2208.12187

29. Enders, A. A., North, N. M., Fensore, C. M., Velez-Alvarez, J. & Allen, H. C. Functional group identification for FTIR spectra using image-based machine learning models. Anal. Chem. 93, 9711–9718 (2021).

30. Zhang, F. & Huang, Q. Characterization of Z-DNA by infrared spectroscopy. Methods Mol. Biol. 2651, 53–58 (2023).

31. Vázquez-Zapién, G. J., Mata-Miranda, M. M., Sánchez-Monroy, V., Delgado-Macuil, R. J., Pérez-Ishiwara, D. G. & Rojas-López, M. FTIR spectroscopic and molecular analysis during differentiation of pluripotent stem cells to pancreatic cells. Stem Cells Int. 2016, 6709714 (2016).

32. Movasaghi, Z., Rehman, S. & Rehman, I. U. Fourier transform infrared (FTIR) spectroscopy of biological tissues. Appl. Spectrosc. Rev. 43, 134–179 (2008).

33. Nogueira, M. S. et al. FTIR spectroscopy as a point-of-care diagnostic tool for diabetes and periodontitis: a saliva analysis approach. Photodiagn. Photodyn. Ther. 40, 103036 (2022).

34. Ye, S., Ruan, P., Yong, J. et al. The impact of the HbA1c level of type 2 diabetics on the structure of haemoglobin. Sci. Rep. 6, 33352 (2016).

35. Vázquez-Zapién, G. J., Mata-Miranda, M. M., Sánchez-Monroy, V., Delgado-Macuil, R. J., Pérez-Ishiwara, D. G. & Rojas-López, M. FTIR spectroscopic and molecular analysis during differentiation of pluripotent stem cells to pancreatic cells. Stem Cells Int. 2016, 6709714 (2016).

36. Arif, B., Ashraf, J. M., Moinuddin, Ahmad, J., Arif, Z. & Alam, K. Structural and immunological characterization of Amadori-rich human serum albumin: role in diabetes mellitus. Arch. Biochem. Biophys. 522, 17–25 (2012).

37. Çimen, D. & Denizli, A. Development of a rapid, sensitive, and effective plasmonic nanosensor for the detection of vitamins in infant formula and milk samples. Photon. Sens. 10, 10.1007/s13320-020-0578-1 (2020).

38. Vázquez-Zapién, G. J., Mata-Miranda, M. M., Sánchez-Monroy, V., Delgado-Macuil, R. J., Pérez-Ishiwara, D. G. & Rojas-López, M. FTIR spectroscopic and molecular analysis during differentiation of pluripotent stem cells to pancreatic cells. Stem Cells Int. 2016, 6709714 (2016).

39. Roy, S. et al. Simultaneous ATR-FTIR-based determination of malaria parasitemia, glucose, and urea in whole blood dried onto a glass slide. Anal. Chem. 89, 5238–5245 (2017).

40. Araújo, R., Ramalhete, L., Ribeiro, E. & Calado, C. Plasma versus serum analysis by FTIR spectroscopy to capture the human physiological state. BioTech 11, 56 (2022).

41. Abe, M., Kitagawa, T. & Kyogoku, Y. Resonance Raman spectra of octaethylporphyrinatoNi(II) and mesodeuterated and 15N-substituted derivatives: II. A normal coordinate analysis. J. Chem. Phys. 69, 4526–4534 (1978).

42. Ghimire, H., Venkataramani, M., Bian, Z., Liu, Y. & Perera, A. G. U. ATR-FTIR spectral discrimination between normal and tumorous mouse models of lymphoma and melanoma from serum samples. Sci. Rep. 7, 16993 (2017).

43. Pakbin, B., Zolghadr, L., Rafiei, S. et al. FTIR differentiation based on genomic DNA for species identification of Shigella isolates from stool samples. Sci. Rep. 12, 2780 (2022).

44. Takamura, A., Watanabe, K., Akutsu, T. et al. Soft and robust identification of body fluid using Fourier transform infrared spectroscopy and chemometric strategies for forensic analysis. Sci. Rep. 8, 8459 (2018).

45. Wang, H. et al. Application of ATR-FTIR spectroscopy and chemometrics for the forensic discrimination of aged peripheral and menstrual bloodstains. Microchem. J. 197, 109933 (2024).

46. Balan, V. et al. Vibrational spectroscopy fingerprinting in medicine: from molecular to clinical practice. Materials 12, 2884 (2019).

47. Sukprasert, J. et al. Synchrotron FTIR light reveals signal changes of biofunctionalized magnetic nanoparticle attachment on Salmonella sp. J. Nanomater. 2020, 6149713 (2020).

48. Sharma, C. P., Sharma, S. & Singh, R. Species discrimination from blood traces using ATR-FTIR spectroscopy and chemometrics: application in wildlife forensics. Forensic Sci. Int. Anim. Environ. 3, 100060 (2023).

49. de Souza, A. T. B. et al. Spectrochemical differentiation in endometriosis based on infrared spectroscopy, advanced data fusion and multivariate analysis. Sci. Rep. 15, 5071 (2025).

50. Scott, D. A. et al. Diabetes-related molecular signatures in infrared spectra of human saliva. Diabetol. Metab. Syndr. 2, 48 (2010).

51. Galant, N. et al. Application of Fourier transform infrared spectroscopy in liquid biopsy to predict the response to first-line immunotherapy in non-small-cell lung cancer patients. Biochem. Biophys. Res. Commun. 771, 152007 (2025).

52. Correia, M. et al. FTIR spectroscopy: a potential tool to identify metabolic changes in dementia patients. J. Alzheimers Neurodegener. Dis. 2, 100007 (2016).

53. Gąsior-Głogowska, M. et al. Spectroscopic techniques in the study of human tissues and their components. Part I: IR spectroscopy. Acta Bioeng. Biomech. 14, 101 (2012).

54. Quinn, A. A. & Elkins, K. M. The differentiation of menstrual from venous blood and other body fluids on various substrates using ATR FT-IR spectroscopy. J. Forensic Sci. 62, 197–204 (2017).

55. Scaggion, C., Marinato, M., Dal Sasso, G. et al. A fresh perspective on infrared spectroscopy as a prescreening method for molecular and stable isotope analyses on ancient human bones. Sci. Rep. 14, 1028 (2024).

56. Alhazmi, H. A. FT-IR spectroscopy for the identification of binding sites and measurements of the binding interactions of important metal ions with bovine serum albumin. Sci. Pharm. 87, 5 (2019).

57. Raouf, G. A., Al-Malki, A.-R. L., Mansouri, N. & Mahmoudi, R. M. Preliminary study in diagnosis and early prediction of preeclampsia by using FTIR spectroscopy technique. J. Am. Sci. 7, 651–659 (2011).

58. Delrue, C., De Bruyne, S. & Speeckaert, M. M. The promise of infrared spectroscopy in liquid biopsies for solid cancer detection. Diagnostics 15, 368 (2025).

59. Mateus, T. et al. Fourier-transform infrared spectroscopy as a discriminatory tool for myotonic dystrophy type 1 metabolism: a pilot study. Int. J. Environ. Res. Public Health 18, 3800 (2021).

60. Abdelrazzak, B. B., Hezma, A. M. & El-Bahy, G. S. ATR-FTIR spectroscopy probing of structural alterations in the cellular membrane of abscopal liver cells. Biochim. Biophys. Acta Biomembr. 1863, 183726 (2021).

61. Wang, X. et al. A study of Parkinson’s disease patients’ serum using FTIR spectroscopy. Infrared Phys. Technol. 106, 103279 (2020).

62. Hano, H., Suarez, B., Lawrie, C. H. & Seifert, A. Fusion of Raman and FTIR spectroscopy data uncovers physiological changes associated with lung cancer. Int. J. Mol. Sci. 25, 10936 (2024).

63. Pang, N. et al. The utilization of blood serum ATR-FTIR spectroscopy for the identification of gastric cancer. Discov. Oncol. 15, 350 (2024).

64. Gąsior-Głogowska, M. et al. Spectroscopic techniques in the study of human tissues and their components. Part I: IR spectroscopy. Acta Bioeng. Biomech. 14, 101 (2012).

65. Thermo Fisher Scientific. ATR-FTIR spectroscopy for forensic tracing: blood stain age (Application Note AN 56372). Thermo Fisher Scientific (n.d.).

66. Stani, C., Vaccari, L., Mitri, E. & Birarda, G. FTIR investigation of the secondary structure of type I collagen: new insight into the amide III band. Spectrochim. Acta A Mol. Biomol. Spectrosc. 229, 118006 (2020).

67. Zupančič, B. et al. Exploration of macromolecular phenotype of human skeletal muscle in diabetes using infrared spectroscopy. Front. Endocrinol. 14, 1308373 (2023).

68. Sharma, C. P., Sharma, S. & Singh, R. Species discrimination from blood traces using ATR-FTIR spectroscopy and chemometrics: application in wildlife forensics. Forensic Sci. Int. Anim. Environ. 3, 100060 (2023).

69. Wang, J. S. et al. FT-IR spectroscopic analysis of normal and cancerous tissues of esophagus. World J. Gastroenterol. 9, 1897–1899 (2003).

70. Ami, D. et al. Fourier transform infrared spectroscopy as a method to study lipid accumulation in oleaginous yeasts. Biotechnol. Biofuels 7, 12 (2014).

71. Ezzati Ghadi, F., Malhotra, A., Ramzani Ghara, A. & Dhawan, D. Modulation of Fourier transform infrared spectra and total sialic acid levels by selenium during 1,2-dimethylhydrazine-induced colon carcinogenesis in rats. Nutr. Cancer 65, 92–98 (2013).

72. de Campos Vidal, B. & Mello, M. L. S. Collagen type I amide I band infrared spectroscopy. Micron 42, 283–289 (2011).

73. Synytsya, A., Vaňková, A., Miškovičová, M., Petrtýl, J. & Petruželka, L. Ex vivo vibration spectroscopic analysis of colorectal polyps for the early diagnosis of colorectal carcinoma. Diagnostics 11, 2048 (2021).

74. Bujok, J. et al. Applicability of FTIR-ATR method to measure carbonyls in blood plasma after physical and mental stress. Biomed. Res. Int. 2019, 2181370 (2019).

75. Olsztyńska-Janus, S., Gąsior-Głogowska, M., Szymborska-Małek, K., Czarnik-Matusewicz, B. & Komorowska, M. Specific applications of vibrational spectroscopy in biomedical engineering. In Biomedical Engineering: Trends, Research and Technologies (ed. Olsztyńska, S.) (InTech, 2011).

76. Scaggion, C., Marinato, M., Dal Sasso, G. et al. A fresh perspective on infrared spectroscopy as a prescreening method for molecular and stable isotope analyses on ancient human bones. Sci. Rep. 14, 1028 (2024).

77. Drobota, M., Grierosu, I., Radu, I. & Stefanescu, C. Modification of protein conformation can be monitored by Fourier transform infrared (FTIR) spectroscopy in oncological patients. Rev. Roum. Chim. 59, 511–515 (2014).

78. Kumar, V., Vora, S., Asodiya, F., Kumar, N. & Gangwar, A. Fourier transform infrared spectroscopy of animal tissues. In Fourier Transform Infrared Spectroscopy (InTechOpen, 2021).

79. Wang, Q. et al. UV–Vis and ATR-FTIR spectroscopic investigations of postmortem interval based on the changes in rabbit plasma. PLoS One 12, e0182161 (2017).

80. Balan, V. et al. Vibrational spectroscopy fingerprinting in medicine: from molecular to clinical practice. Materials 12, 2884 (2019).

81. Sinaei, N., Zare, D. & Azin, M. Production and characterization of poly(3-hydroxybutyrate-co-3-hydroxyvalerate) in wheat starch wastewater and its potential for nanoparticle synthesis. Braz. J. Microbiol. 52, 10.1007/s42770-021-00430-5 (2021).

82. Guo, S. et al. Fast and deep diagnosis using blood-based ATR-FTIR spectroscopy for digestive tract cancers. Biomolecules 12, 1815 (2022).

83. Fadlelmoula, A., Pinho, D., Carvalho, V. H., Catarino, S. O. & Minas, G. Fourier transform infrared (FTIR) spectroscopy to analyse human blood over the last 20 years: a review towards lab-on-a-chip devices. Micromachines 13, 187 (2022).

84. Silva, L. G., Péres, A. F. S., Freitas, D. L. D. et al. ATR-FTIR spectroscopy in blood plasma combined with multivariate analysis to detect HIV infection in pregnant women. Sci. Rep. 10, 20156 (2020).

85. Usoltsev, D., Sitnikova, V., Kajava, A. & Uspenskaya, M. Systematic FTIR spectroscopy study of the secondary structure changes in human serum albumin under various denaturation conditions. Biomolecules 9, 359 (2019).

86. Lin, M. et al. Fast screening of tuberculosis patients based on analysis of plasma by infrared spectroscopy coupled with machine learning approaches. ACS Omega 10, 11817–11827 (2025).

87. Martinez-Cuazitl, A., Vazquez-Zapien, G. J., Sanchez-Brito, M. et al. ATR-FTIR spectrum analysis of saliva samples from COVID-19 positive patients. Sci. Rep. 11, 19980 (2021).

88. Olsztyńska-Janus, S., Gąsior-Głogowska, M., Szymborska-Małek, K., Czarnik-Matusewicz, B. & Komorowska, M. Specific applications of vibrational spectroscopy in biomedical engineering. In Biomedical Engineering: Trends, Research and Technologies (ed. Olsztyńska, S.) (InTech, 2011).

89. Hackett, M. J. et al. Elemental and spectroscopic analysis of ischemic stroke brain tissue using infrared and X-ray microscopy. Anal. Chem. 88, 10949–10956 (2016).

90. Al-Kelani, M. & Buthelezi, N. Advancements in medical research: exploring Fourier transform infrared (FTIR) spectroscopy for tissue, cell, and hair sample analysis. Skin Res. Technol. 30, e13733 (2024).

91. Azimzadeh Andarabi, E., Norouzian-Alam, S., Shayganmanesh, M. & Haji Abdolvahab, M. Analysis of glucose concentrations in blood solutions using FTIR and Raman spectroscopy methods. Biomed. Opt. Express 16, 2631–2662 (2025).

92. Movasaghi, Z., Rehman, S. & Rehman, I. U. Fourier transform infrared (FTIR) spectroscopy of biological tissues. Appl. Spectrosc. Rev. 43, 134–179 (2008).

93. Nogueira, M. S. et al. FTIR spectroscopy as a diagnostic tool for diabetes and periodontitis. Photodiagn. Photodyn. Ther. 40, 103036 (2022).

94. Stani, C., Vaccari, L., Mitri, E. & Birarda, G. FTIR investigation of type I collagen amide III band. Spectrochim. Acta A Mol. Biomol. Spectrosc. 229, 118006 (2020).

95. Sharma, C. P., Sharma, S. & Singh, R. Species discrimination from blood traces using ATR-FTIR spectroscopy and chemometrics: application in wildlife forensics. Forensic Sci. Int. Anim. Environ. 3, 100060 (2023).

96. Kong, J. & Yu, S. Fourier transform infrared spectroscopic analysis of protein secondary structures. Acta Biochim. Biophys. Sin. 39, 549–559 (2007).

97. Wang, X. et al. Serum FTIR spectroscopy in Parkinson’s disease. Infrared Phys. Technol. 106, 103279 (2020).

98. Takamura, A., Watanabe, K., Akutsu, T. et al. FTIR spectroscopy for forensic identification of body fluids. Sci. Rep. 8, 8459 (2018).

99. Kong, J. & Yu, S. Fourier transform infrared spectroscopic analysis of protein secondary structures. Acta Biochim. Biophys. Sin. 39, 549–559 (2007).

100. Wang, X. et al. Serum FTIR spectroscopy in Parkinson’s disease. Infrared Phys. Technol. 106, 103279 (2020).

101. Takamura, A., Watanabe, K., Akutsu, T. et al. FTIR spectroscopy for forensic identification of body fluids. Sci. Rep. 8, 8459 (2018).

102. Ye, S., Ruan, P., Yong, J., Shen, H., Liao, Z. & Dong, X. The impact of the HbA1c level of type 2 diabetics on the structure of haemoglobin. Sci. Rep. 6, 33352 (2016).

103. Hamieda, S., Hassan, A. & Saied, M. Biophysical and biochemical study exploring resveratrol protective potential against gamma irradiation effects. Bull. Natl. Res. Cent. 48, 1280 (2024).

104. Azimzadeh Andarabi, E., Norouzian-Alam, S., Shayganmanesh, M. & Haji Abdolvahab, M. Analysis of glucose concentrations in blood solutions using FTIR and Raman spectroscopy methods. Biomed. Opt. Express 16, 2631–2662 (2025).

105. McAvan, B. S., France, A. P., Bellina, B., Barran, P. E., Goodacre, R. & Doig, A. J. Quantification of protein glycation using vibrational spectroscopy. Analyst 145, 3686–3696 (2020).

106. Ioannou, A. & Varotsis, C. Modifications of hemoglobin and myoglobin by Maillard reaction products (MRPs). PLoS One 12, e0188095 (2017).

107. Baldwin, J. & Chothia, C. Haemoglobin: the structural changes related to ligand binding and its allosteric mechanism. J. Mol. Biol. 129, 175–220 (1979).

108. Huang, Y.-T., Liao, H.-F., Wang, S.-L. & Lin, S.-Y. Glycation and secondary conformational changes of human serum albumin: study of the FTIR spectroscopic curve-fitting technique. AIMS Biophys. 3, 247–260 (2016).

109. Barth, A. & Zscherp, C. What vibrations tell us about proteins. Q. Rev. Biophys. 35, 369–430 (2002).

110. Baker, M. J. et al. Using Fourier transform IR spectroscopy to analyze biological materials. Nat. Protoc. 9, 1771–1791 (2014).

111. Movasaghi, Z., Rehman, S. & Rehman, I. U. Fourier transform infrared (FTIR) spectroscopy of biological tissues. Appl. Spectrosc. Rev. 43, 134–179 (2008).

112. Wold, S., Sjöström, M. & Eriksson, L. PLS-regression: a basic tool of chemometrics. Chemom. Intell. Lab. Syst. 58, 109–130 (2001).

113. Næs, T., Isaksson, T., Fearn, T. & Davies, T. A User-Friendly Guide to Multivariate Calibration and Classification (NIR Publications, 2002).

114. Kauppinen, J. K., Moffatt, D. J., Mantsch, H. H. & Cameron, D. G. Fourier self-deconvolution: a method for resolving intrinsically overlapped bands. Appl. Spectrosc. 35, 271–276 (1981).

115. de Juan, A. & Tauler, R. Chemometrics applied to unravel multicomponent processes and mixtures: revisiting latest trends in multivariate resolution. Anal. Chim. Acta 500, 195–210(2003).

116. Krizhevsky, A., Sutskever, I. & Hinton, G. E. ImageNet classification with deep convolutional neural networks. Commun. ACM 60, 84–90 (2017).

117. Liu, Y., Chen, Y., Dong, L. et al. Deep learning in infrared spectroscopy: a review. TrAC Trends Anal. Chem. 120, 115652 (2019).

118. Acquarelli, J., van Laarhoven, T., Gerretzen, J. et al. Convolutional neural networks for vibrational spectroscopic data analysis. Anal. Chim. Acta 954, 22–31 (2017).

119. Lundberg, S. M. & Lee, S. I. A unified approach to interpreting model predictions. Adv. Neural Inf. Process. Syst. 30, 4765–4774 (2017).

120. Arefi, A., Sturm, B. & Hoffmann, T. Explainability of deep convolutional neural networks when it comes to NIR spectral data: a case study of starch content estimation in potato tubers. Food Control 169, (2025).

121. Antonio, D., O’Toole, H., Carney, R., Kulkarni, A. & Palazoglu, A. Assessing the performance of 1D-convolution neural networks to predict concentration of mixture components from Raman spectra. arXiv preprint arXiv:2306.16621 (2023).

122. Chen, R., et al. Enhancing glucose analysis using FTIR-ATR spectroscopy: comprehensive pre-processing methods and multi-feature values-based machine learning. SSRN Electron. J. (2024). Available at SSRN: https://ssrn.com/abstract=4936430.

123. Mishra, S., Prasad, A. K., Vinod, A. et al. A multi-model approach for estimation of ash yield in coal using Fourier transform infrared spectroscopy. Sci. Rep. 15, 13786 (2025).

124. Bioucas-Dias, J. M. et al. Hyperspectral remote sensing data analysis and future challenges. IEEE Geosci. Remote Sens. Mag. 1, 6–36 (2013).

