## Supplementary Information for "Decoupling Accuracy and Explainability: Machine Learning Strategies for HbA1c Prediction and Biomarker Discovery in Blood FTIR Spectroscopy"

#### Data preparation

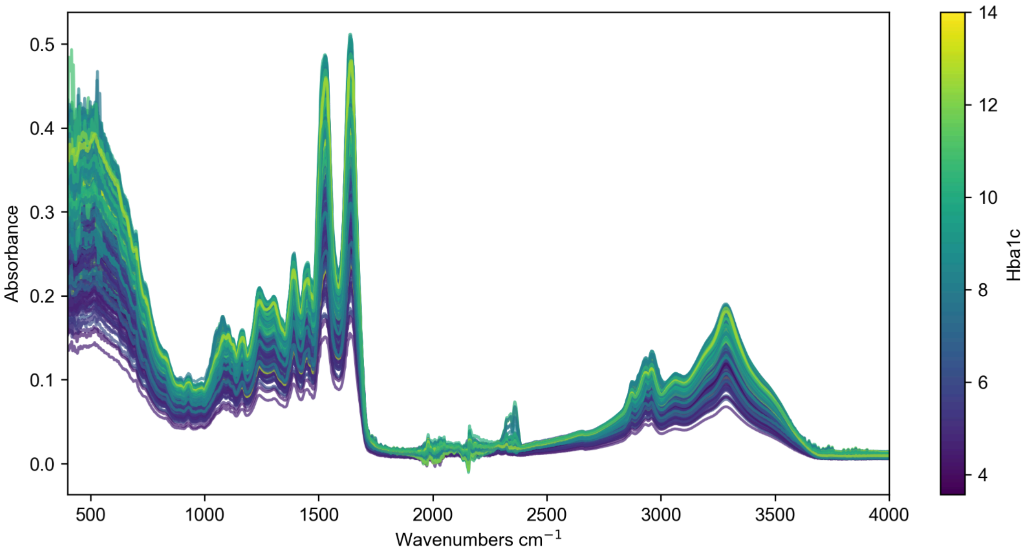


**Figure 1** FTIR absorbance spectra of blood samples across the full mid-infrared range (400–4000 cm⁻¹). Each line represents an individual sample, color-coded according to HbA1c level (scale bar on the right, ranging from ~4% to 14%). Major absorbance features are evident in the bio-fingerprint

region (1000–1800 cm⁻¹), CH-stretching region (2800–3000 cm⁻¹), and amide/O–H region (~3300 cm⁻¹). The overlay highlights spectral variability between samples while preserving consistent band positions, with intensity differences reflecting underlying biochemical variation related to HbA1c.

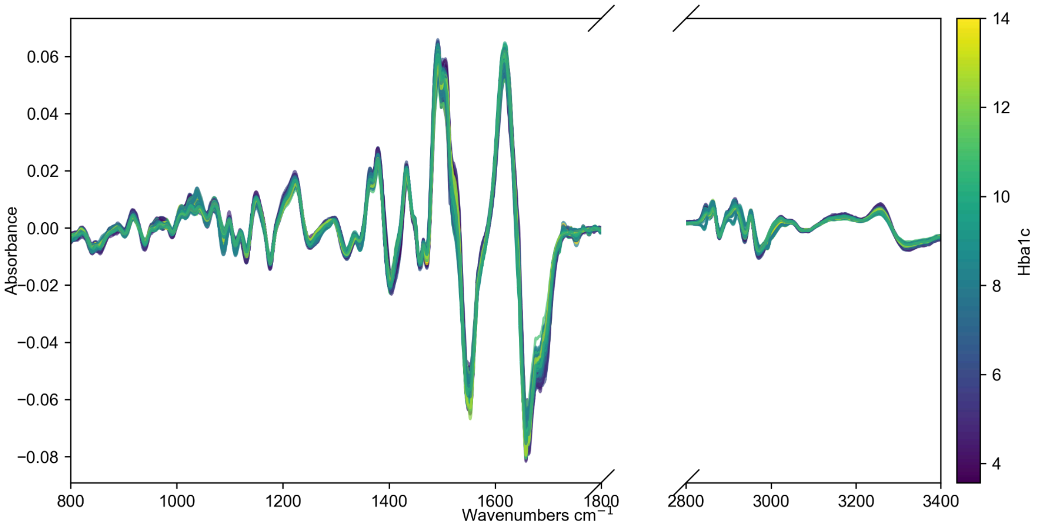


**Figure 2.** Preprocessed FTIR spectra prepared for PLSR/CNN modeling. Signal processing includes a Savitzky-Golay filter (1st derivative) to resolve overlapping peaks and remove baseline offsets, followed by vector normalization to correct for sample thickness variations. The noisy low-wavenumber region (400–800 $cm^{-1}$) and the non-informative silent region (~1800–2800 $cm^{-1}$) were cut to isolate relevant biological features and enhance signal quality.

### Curve Fitting

The pseudo-Voigt function approximates a Voigt profile through a linear combination of Gaussian and Lorentzian profiles, weighted by a mixing parameter $\eta$ where $0 < \eta < 1$. The general form is defined as:

$$V(x, x_0, A, \Gamma, \eta) = A[\eta G(x, x_0, \Gamma) + (1 - \eta) L(x, x_0, \Gamma)]$$

Here, $x_0$ represents the center of the distribution, $A$ is the area, and $\Gamma$ is the full width at half maximum (FWHM). which indicates the width of the spectral curve measured between two points at half of the maximum height. Additional parameters included height, which is the highest value of the peak.

The Gaussian component, $G(x, x_0, \Gamma)$, is normalized to unit area and expressed as:

$$G(x, x_0, \Gamma) = a_G e^{-b_G (x - x_0)^2}$$

The constants are derived from the FWHM such that:

$$\text{Where } a_G = \frac{2}{\Gamma}\sqrt{\frac{\ln 2}{\pi}} \text{ and } b_G = \frac{4\ln 2}{\Gamma^2}$$

The Lorentzian component, $L(x,\ x_0,\ \Gamma)$, is similarly normalized and defined as:

$$L(x,\ x_0,\ \Gamma) = \frac{1}{\pi}\frac{\Gamma/2}{(x - x_0)^2 + (\Gamma/2)^2}$$

Since the resulting objective function to fit is non-convex, numerous local minima can yield similar goodness-of-fit metrics but may not necessarily reflect the biologically correct solution. To address this issue some of the starting parameters were fixed. Specifically, peak centers were identified using the second derivative of the spectra and used to initialize the $x_0$ parameter, allowing for a variation of ±3 $cm^{-1}$ around the center. Areas were constrained to positive values, with the initial amplitude set to the absorbance at the peak center of the preprocessed spectrum. The full width at half maximum (FWHM) was also restricted to positive values, initialized arbitrarily at 10. The mixing parameter was set to vary within the range (0, 1), with an initial value of 0.5.

Peak detection was performed using the following automated algorithm:

1) The spectrum was divided into two regions: 1000–1520 $cm^{-1}$ and 1480–1720 $cm^{-1}$. To minimize artifacts from spectral smoothing near the boundaries, additional data beyond the region edges were incorporated during processing, enhancing the robustness of the analysis.
2) The second derivative of all spectra was computed using the Savitzky-Golay filter, applying region-specific smoothing parameters to address varying signal-to-noise ratios: window size of 20, polynomial order of 2 for the first region, and window size of 5, polynomial order of 2 for the second region.
3) Peaks were identified using scipy.signal.find_peaks. To account for minor peak shifting (which may manifest as distinct features across the dataset), only peaks occurring with a frequency of at least 10% across all spectra were retained for the final model (Fig. 3).

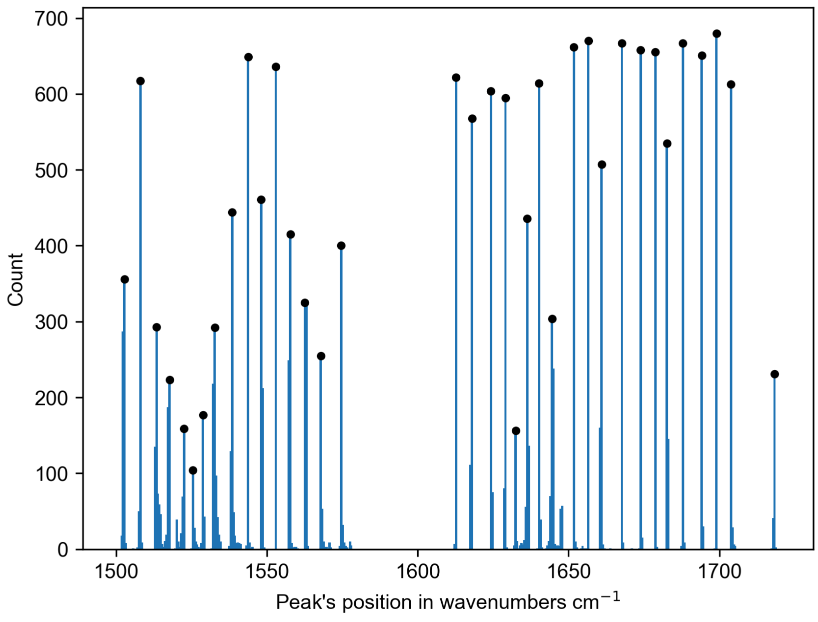


**Figure 3** *Distribution of detected peaks in the amide I and II regions of the spectra. Each bar represents the frequency of a peak detected at a given wavenumber, and black markers indicate peaks that occurred in at least 10% of samples. The plot highlights recurrent features across the dataset, with consistently observed bands concentrated in the 1500–1720 cm$^{-1}$ range.*

#### Models quality evaluation

RMSE (Root Mean Squared Error) measures the standard deviation of the prediction errors. It is calculated as:

$$RMSE = \sqrt{\frac{1}{n}\sum_{i=1}^{n}(y_i - \hat{y}_i)^2}$$

Where $y_i$ are the actual values, $\hat{y}_i$ are the predicted values, and $n$ is the number of samples. In this work, this metric is denoted as RMSECV when derived from cross-validation and RMSEP when evaluated on out-of-sample data.

MSE is a common metric used to quantify the difference between predicted and actual values. It is calculated as the average of the squared differences between predicted and actual HbA1c values:

$$MSE = \frac{1}{n}\sum_{i=1}^{n}(y_i - \hat{y}_i)^2$$

MSE penalizes larger errors more heavily, making it particularly sensitive to outliers. A lower MSE value indicates a better fit of the model to the data. Since MSE was chosen as the loss function for the CNN model, the terms *MSE* and *loss function* are used interchangeably in this context.

MAE quantifies how close predictions are to the actual values by calculating the average absolute difference. MAE measures how accurately a model predicts outputs. A smaller MAE indicates better model performance.

$$MAE = \frac{1}{n}\sum_{i=1}^{n} |y_i - \hat{y}_i|$$

The $R^2$ value, also known as the coefficient of determination, represents the proportion of variance in the actual HbA1c values that is explained by the model. It is calculated as:

$$R^2 = 1 - \frac{RSS}{TSS}$$

Where is the mean of the actual values. $R^2$ ranges from 0 to 1, where a value of 1 indicates that the model perfectly explains the variance in the actual values, and a value close to 0 suggests that the model has little explanatory power.

Discrepancy (DIS) was used to assess the goodness of fit for the fitted spectrum. While DIS is essentially equivalent to RMSE, we retain this terminology to distinguish between the goodness of fit in curve fitting and the evaluation of model performance.
